# Quantifying the symptom burden of COVID-19: pre-infection through 1 month

**DOI:** 10.64898/2026.08.07.26359811

**Authors:** Alon Yehoshua, Laura L. Lupton, Tianyan Hu, Joseph C. Cappelleri, Meghan B. Gavaghan, Laura Puzniak, Rachel Brathwaite, Manuela Di Fusco, Xiaowu Sun

## Abstract

**Background:** To characterize Coronavirus disease 2019 (COVID-19) symptom severity, and recovery from pre-infection through one month, overall and by risk groups.

**Methods:** Symptomatic adults aged ≥18 years with test-confirmed COVID-19 were enrolled from ambulatory care clinics within a national U.S. retail pharmacy network between 10/24/2024 and 08/29/2025 (NCT05160636). Adjusted mixed models for repeated measures estimated least-squares mean changes (LSE) and standard errors (SE) from pre-infection and on Days 1-7, 10, 14, and Week 4 from enrollment in composite symptom scores (sum of severity ratings (0-3) across 14 symptoms), counts of mild-to-severe, moderate-to-severe, and severe symptoms, overall and by age and clinical risk status. Effect sizes (ES) were defined as small (0.2-<0.5), medium (≥0.5), and large (≥0.8).

**Results:** The analysis included 608 adults. On Day 1, symptom severity rose sharply from pre-infection for the composite symptom score (LSE 14.2 [SE 0.3]; ES 2.22), mild-to-severe (7.6 [0.1]; 2.72), moderate-to-severe (5.0 [0.2]; 1.77); and severe (1.8 [0.1]; 0.92) (all p<0.001). By Week 4, composite score (0.7 [0.2]; 0.26), mild-to-severe (0.5 [0.1]; 0.23); moderate-to-severe symptoms (0.1 [0.1]; 0.17) and severe symptoms (0.2 [0.1]; 0.5) remained slightly above baseline (all p≤0.025). Elevated severe symptom durations varied: high-risk adults (through Day 3), adults <50 years (through Day 7), and adults ≥50 years (through Day 7).

**Conclusions:** COVID-19 was associated with notable acute symptoms in outpatients, followed by gradual improvement over time, although symptoms still persisted at four weeks. Improvement in severe symptoms varied by individual risk profile, reinforcing the importance risk-based follow-up and ongoing monitoring.

## INTRODUCTION

Since its emergence in late 2019, COVID-19 has resulted in widespread infection and remains an ongoing public health concern. Hospitalization and mortality rates from SARS-CoV-2 have decreased since the early pandemic [1]. However, ongoing circulation of the SARS-CoV-2 virus continues to pose a significant clinical and public health challenge, especially in outpatient and community settings [2,3].

Clinical manifestations of COVID-19 range from asymptomatic infection to symptomatic illness of varying severity. Among symptomatic individuals, illness severity and recovery are influenced by factors such as vaccination status, age, and the presence of underlying medical conditions [4,5]. COVID-19 symptoms commonly involve respiratory, systemic, and gastrointestinal symptoms and may vary in both intensity and duration. While individuals of all ages are affected, older adults and those with comorbid conditions remain at increased risk for adverse clinical outcomes [6,7].

Despite the established associations between age, comorbidities, and risk for severe COVID-19 outcomes, there remains limited evidence characterizing the nature of symptom burden changes from the pre-infection period through recovery among outpatients overall and among high-risk groups [8]. This characterization is clinically important to distinguish symptom burden attributable to acute COVID-19 from baseline health status especially among different risk groups with pre-existing comorbidities and established clinical risk of severe disease.

Prior COVID-19 burden studies using retail pharmacy recruitment primarily summarized symptoms using overall prevalence or symptom counts and evaluated changes at the aggregate level [11–13]. These studies did not characterize symptom burden at the individual-symptom level or evaluate changes in the symptom presence and severity of specific symptoms over time. To address these evidence gaps, this real-world outpatient study was performed to provide a more granular assessment of symptom burden by characterizing the changes in the presence and severity of 14 COVID-19-related symptoms from pre-infection period through one month following positive SARS-COV-2 test among symptomatic adults. In addition, symptom burden and trajectories are described among high-risk subgroups, such as older adults and individuals with comorbid conditions, to better characterize any differences in symptom trajectory during the first month following infection.

## METHODS

### Study design and participants

This prospective, repeated-measures study enrolled symptomatic adults (≥18 years) with test-confirmed COVID-19 infection from ambulatory care clinics within a nationwide US retail pharmacy chain (10/24/2024 to 08/29/2025). Eligible individuals were invited by email within 48 hours of a positive rapid antigen or molecular test and completed electronic consent.

Inclusion criteria required symptoms beginning on or within four days before testing, English proficiency, and internet access. Exclusion criteria included recent COVID-19 vaccination (within 14 days prior to symptom onset), recent SARS-CoV-2 or influenza infection (within 30 days prior to completing the screening questionnaire), receipt of a non-Pfizer BioNTech vaccine after authorization of the updated 2024-2025 formulation on 08/22/2024 [14], vaccinated prior to 08/23/2024 and had received their most recent COVID-19 vaccine within 6 months of the positive test date. All participants received compensation for completing surveys.

### Sociodemographic and clinical variables

Participants self-reported demographics, insurance type and employment status, COVID-19 vaccination, antiviral use, and CDC-defined risk conditions [4]. Social Vulnerability Index (SVI) was derived from ZIP codes (scores ranged from 0-1, higher scores represent more vulnerable communities) [15]. High-risk status was defined as adults aged ≥65 years or adults of any age with ≥1 CDC-identified COVID-19 risk condition or pregnancy [4] (See Supporting Information).

### Assessment of symptom burden

Participants rated the presence and severity of 14 CDC-identified acute COVID-19-related symptoms [16]. These included systemic (feeling feverish, chills or shivering, muscle or body aches, headache, fatigue), respiratory (shortness of breath, nasal congestion or runny nose, cough, sore throat), gastrointestinal (nausea, vomiting, diarrhea), and neurological symptoms (new loss of taste or smell). Level of severity of systemic, respiratory, and nausea symptoms were rated on a 4-point ordinal scale (0=None, 1=Mild, 2=Moderate, 3=Severe), while the frequency of vomiting and diarrhea in the last 24 hours was reported (0=0 times, 1=1-2 times, 2=3-4 times, or 3=5 or more times). Changes in taste/smell in the last 24 hours were coded as 0=No change, 1=Change, 2=Cannot smell/taste). On Day 1, participants retrospectively rated symptoms for pre-infection, the worst point of current infection from symptom onset to study enrollment, and the worst point during the past 24 hours. Participants subsequently completed symptom surveys on Days 2-7, 10, 14, and Week 4 from study enrollment to capture symptom presence and severity.

Symptom burden was assessed using four outcomes: 1) a composite score summing the ratings from 14 symptoms [17]; 2) count of mild-to-severe symptoms; 3) count of moderate-to-severe symptoms; 4) count of severe symptoms. Higher scores indicated greater burden.

### Statistical analysis

Categorical variables were summarized as counts and percentages, and continuous variables as means and standard deviations (SD). Mixed-effects models for repeated measures (MMRM) estimated least-squares means and mean changes in outcomes with standard errors (SE) [18]. Time was modeled as a categorical variable to allow for non-linear changes in symptom outcomes relative to the worst moment. All models adjusted for: time (categorical: worst moment (reference), Day 1, Day 2, Day 3, Day 4, Day 5, Day 6, Day 7, Day 10, Day 14, and Week 4), clinical high-risk status (high-risk/not high-risk), self-reported sex (male/female), age group (≥50 vs <50), race/ethnicity (White, Black or African American, Hispanic or Latino, Other), SVI (categorical: <0.25, ≥0.25 to <0.5, ≥0.5 to <0.75, ≥0.75), US region (Northeast, South, Midwest, West, Other), number of comorbidities (categorical: 0, 1-2, ≥3)), insurance type, and the baseline score (pre-infection outcome measure). Interaction terms between clinical high-risk status and time, and age group and time were included. Repeated measures were modeled using an unstructured covariance matrix. MMRM model parameters are shown in Table S1 (Supporting Information). Mean changes from pre-infection were evaluated using model-based t-tests (p<0.05, no multiplicity adjustment). Within-cohort (standardized) effect size (ES) was calculated by dividing the mean change from pre-infection to post-baseline by the SD of the score change. ES were interpreted using Cohen’s d thresholds: small (0.2 - <0.5), medium (0.5 - <0.8), or large (≥0.8) [19,20]. Visual summaries were generated to illustrate symptom trajectories over time.

MMRM analyses were conducted under the assumption that data were missing at random (MAR); no imputation was performed for missing data. This study was reported in accordance with the Strengthening the Reporting of Observational Studies in Epidemiology (STROBE) recommendations [21]. All analyses were conducted in SAS Version 9.4 (SAS Institute, Cary, NC).

### Subgroup analysis by age and underlying health conditions

Prespecified subgroup analyses described symptom burden overall and within three clinically relevant demographic and risk groups: 1) high-risk adults; 2) adults ≥50 years and 3) adults <50 years.

## RESULTS

### Description of baseline sociodemographic and clinical characteristics

A total of 608 adults were included in the analytic sample (Figure 1). The median time from symptom onset to enrollment was 4 days (interquartile range (IQR) = 3-5). The mean age was 45.9 (SD = 14.2) years (Table 1). Most participants were females (76.5%), White (71.7%), and employed (78.3%). Participants predominantly resided in the South (60.5%) in moderately vulnerable communities (mean SVI = 0.37). Overall, 17.4% had received the 2024-2025 Pfizer-BioNTech COVID-19 vaccine, and 53.9% reported antiviral use (which included treatment with either molnupiravir (Lagevrio™), nirmatrelvir/ritonavir (Paxlovid™), or other antiviral medications). A total of 379 participants (62.3%) were considered high-risk, while 244 participants (40.1%) were ≥ 50 years and 364 (59.9%) were <50 years. Obesity (body mass index (BMI) >30) was the most prevalent (30.1%) comorbidity, followed by mental health conditions (23.4%) and smoking (14.5%); five (0.8%) participants were pregnant. Survey response rates declined over follow-up, with 70.1% completing the Week 4 assessment (Table 2).

**Figure 1.**
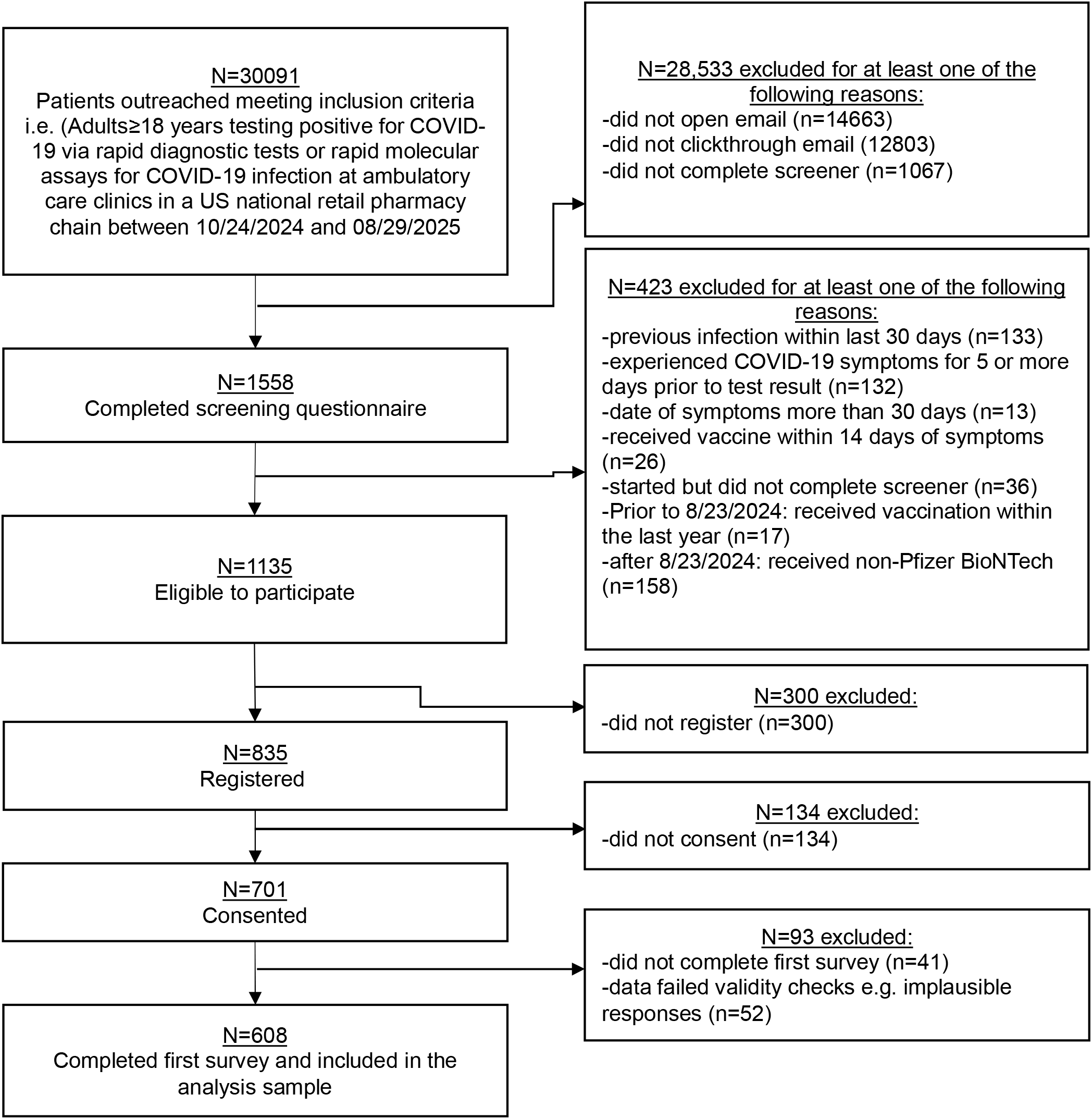
Study participant flow chart

**Table 1.** Baseline characteristics.

|  | **Overall**<br>**sample**<br>**(N=608)** | **COVID-19**<br>**high-risk**<br>**population**<br>**(N=379)†** |
| --- | --- | --- |
| Age, years, mean (SD) | 45.9 (14.2) | 48.5 (14.9) |
| Self-reported sex, n (%) |  |  |
| Female | 465 (76.5) | 295 (77.8) |
| Race/Ethnicity, n (%) |  |  |
| White | 436 (71.7) | 288 (76.0) |
| Black or African American | 48 (7.9) | 33 (8.7) |
| Hispanic or Latino | 79 (13.0) | 36 (9.5) |
| Asian | 25 (4.1) | 11 (2.9) |
| Other | 20 (3.3) | 11 (2.9) |
| US Region of residence, n (%) |  |  |
| Northeast | 73 (12.0) | 42 (11.1) |
| South | 368 (60.5) | 230 (60.7) |
| Midwest | 133 (21.9) | 88 (23.2) |
| West | 33 (5.4) | 19 (5.0) |
| Other\Unknown | 1 (0.2) | (0.0) |
| Employment status, n (%) |  |  |
| Yes | 476 (78.3) | 283 (74.7) |
| Social Vulnerability Index <sup>a</sup>, mean (SD) | 0.37 (0.21) | 0.38 (0.21) |
| Social Vulnerability Index category<sup>‡</sup>, n (%) |  |  |
| <0.25 | 205 (33.7) | 120 (31.7) |
| ≥0.25 and <0.5 | 244 (40.1) | 157 (41.4) |
| ≥0.5 and <0.75 | 129 (21.2) | 81 (21.4) |
| ≥0.75 | 30 (4.9) | 21 (5.5) |
| Antiviral use, n (%) | 328 (53.9) | 226 (59.6) |
| Received 2024-2025 Pfizer-BioNTech vaccine, n (%) | 106 (17.4) | 75 (19.8) |
| Pregnant, n (%) |  |  |
| No | 414 (68.1) | 246 (64.9) |
| Yes | 5 (0.8) | 5 (1.3) |
| Not applicable | 189 (31.1) | 128 (33.8) |
| Underlying health conditions, n (%) |  |  |
| Any current/active cancers or<br>malignancies<sup>§</sup> | 7 (1.2) | 7 (1.8) |
| Cerebrovascular disease | 7 (1.2) | 7 (1.8) |
| Chronic kidney disease | 7 (1.2) | 7 (1.8) |
| Chronic lung conditions | 65 (10.7) | 65 (17.2) |
| Chronic liver disease | 12 (2.0) | 12 (3.2) |
| Endocrine disorders | 51 (8.4) | 51 (13.5) |
| Heart conditions | 22 (3.6) | 22 (5.8) |
| Mental health conditions | 142 (23.4) | 142 (37.5) |
| Obesity: BMI>30 | 183 (30.1) | 183 (48.3) |
| Weakened immune system/<br>immunocompromised | 25 (4.1) | 25 (6.6) |
| Smoker | 88 (14.5) | 88 (23.2) |
| Active tuberculosis | 0 | 0 |
| Number of comorbidities, mean (SD) | 1.0 (1.1) | 1.6 (1.0) |
| ≥1 comorbidity, n (%) | 352 (57.9) | 352 (92.9) |

|  | **Overall**<br>**sample**<br>**(N=608)** | **COVID-19**<br>**high-risk**<br>**population**<br>**(N=379)<sup>†</sup>** |
| --- | --- | --- |
| ≥1 comorbidity or pregnant, n (%) | 354 (58.2) | 354 (93.4) |
| Time from symptom onset to<br>enrollment, days, Median (IQR) | 4 (3~5) | 4 (3~5) |
| Time from vaccination to<br>enrollment, days, Median (IQR) | 964 (431.5-1260) | 890<br>(369~1,230) |

BMI=body mass index; IQR=Interquartile range; SD=standard deviation; US=United States.
<sup>†</sup>High-risk criteria for COVID-19 (defined as adults aged ≥65 years and adults <65 years with ≥1 CDC-recognized underlying health condition associated with increased risk of getting very sick from COVID-19).
<sup>‡</sup>SVI is a score that ranges from 0 to 1. Higher values represent higher vulnerability.
<sup>§</sup>Any current/active cancers or malignancies (other than skin cancer)

**Table 2.** Summary of least-square estimates^†^ of mean change in symptom measures from prior to infection in total population.

| Time | Retention rate<br>n (%) | Composite acute symptom score |  |  | Number of mild to severe acute symptoms |  |  | Number of moderate to severe acute symptoms |  |  | Number of severe acute symptoms |  |  |
| --- | --- | --- | --- | --- | --- | --- | --- | --- | --- | --- | --- | --- | --- |
|  |  | LSE (SE) | ES<sup>‡</sup> | p-value | LSE (SE) | ES<sup>‡</sup> | p-value | LSE (SE) | ES<sup>‡</sup> | p-value | LSE (SE) | ES<sup>‡</sup> | p-value |
| Prior to infection<sup>§</sup> | 608 (100.0) | 1.5 (1.9) |  |  | 1.4 (1.6) |  |  | 0.1 (0.4) |  |  | 0.0 (0.1) |  |  |
| Worst Moment | 608 (100.0) | 17.5 (0.3) | 2.81 | **<0.001** | 8.2 (0.1) | 3.37 | **<0.001** | 6.5 (0.1) | 2.47 | **<0.001** | 3.1 (0.1) | 1.19 | **<0.001** |
| Day 1 | 608 (100.0) | 14.2 (0.3) | 2.22 | **<0.001** | 7.6 (0.1) | 2.72 | **<0.001** | 5.0 (0.1) | 1.77 | **<0.001** | 1.8 (0.1) | 0.92 | **<0.001** |
| Day 2 | 508 (83.6) | 7.6 (0.3) | 1.44 | **<0.001** | 5.1 (0.1) | 1.76 | **<0.001** | 2.1 (0.1) | 0.92 | **<0.001** | 0.6 (0.1) | 0.52 | **<0.001** |
| Day 3 | 516 (84.9) | 5.7 (0.2) | 1.22 | **<0.001** | 4.1 (0.1) | 1.46 | **<0.001** | 1.4 (0.1) | 0.71 | **<0.001** | 0.4 (0.1) | 0.51 | **<0.001** |
| Day 4 | 503 (82.7) | 4.4 (0.2) | 1.10 | **<0.001** | 3.3 (0.1) | 1.27 | **<0.001** | 0.9 (0.1) | 0.58 | **<0.001** | 0.3 (0.1) | 0.46 | **<0.001** |
| Day 5 | 474 (78.0) | 3.3 (0.2) | 0.93 | **<0.001** | 2.6 (0.1) | 1.04 | **<0.001** | 0.6 (0.1) | 0.46 | **<0.001** | 0.2 (0.1) | 0.41 | **<0.001** |
| Day 6 | 483 (79.4) | 3.0 (0.2) | 0.83 | **<0.001** | 2.3 (0.1) | 0.89 | **<0.001** | 0.6 (0.1) | 0.44 | **<0.001** | 0.2 (0.1) | 0.40 | **0.001** |
| Day 7 | 472 (77.6) | 2.4 (0.2) | 0.69 | **<0.001** | 1.9 (0.1) | 0.77 | **<0.001** | 0.5 (0.1) | 0.36 | **<0.001** | 0.2 (0.1) | 0.39 | **0.004** |
| Day 10 | 439 (72.2) | 2.0 (0.2) | 0.66 | **<0.001** | 1.5 (0.1) | 0.68 | **<0.001** | 0.4 (0.1) | 0.38 | **<0.001** | 0.2 (0.1) | 0.43 | **0.013** |
| Day 14 | 432 (71.1) | 1.4 (0.2) | 0.50 | **<0.001** | 1.1 (0.1) | 0.50 | **<0.001** | 0.3 (0.1) | 0.31 | **<0.001** | 0.2 (0.1) | 0.48 | **0.009** |
| Week 4 | 426 (70.1) | 0.8 (0.1) | 0.32 | **<0.001** | 0.6 (0.1) | 0.28 | **<0.001** | 0.178<br>(0.043) | 0.24 | **<0.001** | 0.2 (0.1) | 0.50 | **0.025** |

LSE=least-square estimate; SE=standard error; ES=effect size.
<sup>†</sup>Least-square estimates based on mixed models for repeated measures. See Table S1 for model parameters.
<sup>‡</sup> ES refers to within-cohort effect size, was calculated as the least square estimate of mean change scores divided by the observed standard deviation of change scores from prior to infection to follow-up.
<sup>§</sup> Mean (standard deviation) of scores prior to infection reflect observed values and were not derived using the least squares model estimates.
Bolded values indicate P<0.05 which is considered statistically significant.

### Symptom severity prevalence

On Day 1 (median of 4 days since symptom onset), respiratory symptoms (stuffy or runny nose (97.4%), cough (94.1%)), and systemic symptoms (fatigue or tiredness (93.6%), headache (85.7%), and body ache (81.4%), were highly prevalent (Table S2, and Figure 2A). By Week 4, stuffy or runny nose (35.0%), and cough (30.3%) remained the most prevalent symptoms, followed by fatigue or tiredness (28.9%), body ache (22.8%) and headache (22.1%). These symptoms remained the most reported across the one-month follow-up.

**Figure 2.**
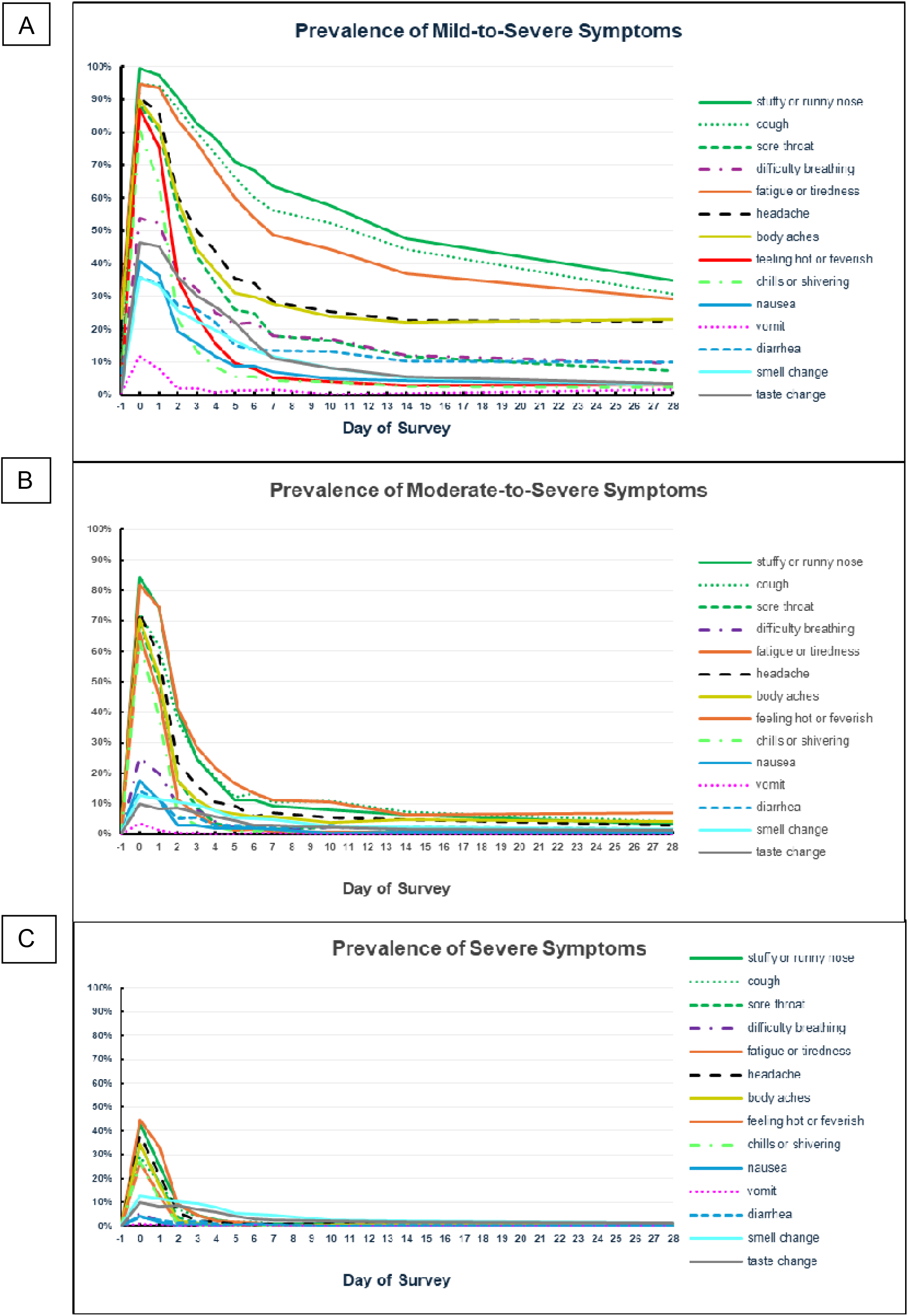
Prevalence of symptoms over 4 weeks: A) Mild-to-Severe; B) Moderate-to-Severe; C) Severe. Day −1 is a placeholder to reflect the period prior to infection or symptom onset; Day 0 is a placeholder to represent the worst moment of illness since infected as reported by participants.

On Day 1, the most common moderate-to-severe symptoms were fatigue or tiredness (74.2%), stuffy or runny nose (73.9%) and cough (61.5%) (Table S2, and Figure 2B), though the prevalence fell sharply by Week 4 (≤7%).

The severe symptoms most reported on Day 1 (median 4 days since symptom onset) were fatigue or tiredness (32.7%), stuffy or runny nose (25.5%) and headache (20.9%). However, these declined rapidly with ≤5% reporting any severe symptom after Day 6. By Week 4, severe symptoms were rare (<1-1.4%) (Table S2, and Figure 2C).

### Mean symptom changes from prior-to-infection through one month post diagnosis

Relative to the pre-infection period, symptom burden increased markedly at Day 1 across all symptom measures (all p<0.001) (Table 2 and Figure 3A-3B). Specifically, the mean composite score increased by 14.2 points (SE: 0.3), accompanied by increases of 7.6 (SE: 0.1) mild-to-severe, 5.0 (SE: 0.1) moderate-to-severe, and 1.8 (SE: 0.1) severe symptoms at Day 1 (Table 2 and Figure 3B). Within-cohort ES for composite score (ES: 2.22), mean number of mild-to-severe (ES: 2.72), moderate-to-severe (ES:1.77) and severe symptoms (ES: 0.92) were large on Day 1.

**Figure 3.**
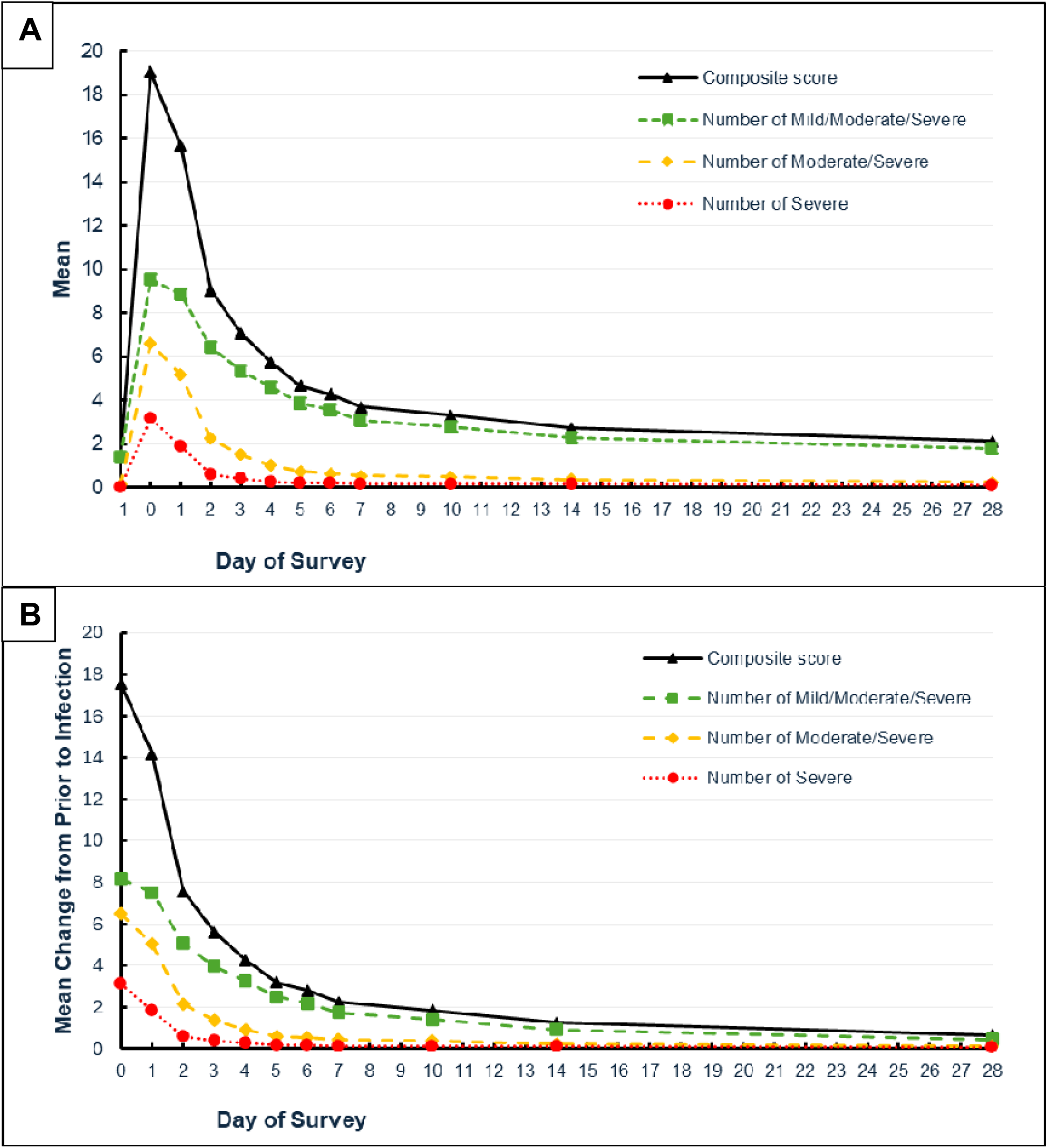
Symptom severity measures over 4 weeks A) mean; B) mean changes from prior to infection. Day −1 is a numerical placeholder to reflect the period prior to infection or symptom onset; Day 0 is a numerical placeholder that represents the worst moment of illness since infected as reported by participants.

Following this initial peak, symptom burden declined steadily across all measures. However, all symptom indices remained elevated relative to pre-infection levels at Week 4. At Week 4, the composite symptom score for the total population remained higher by 0.8 (SE: 0.1; p<0.001), ES: 0.32; the mean number of mild-to-severe symptoms by 0.6 (SE: 0.1; p<0.001), ES: 0.28; the mean number of moderate-to-severe symptoms by 0.2 (SE: 0.004; p<0.001), ES: 0.24; and the mean number of severe symptoms by 0.2 (SE:0.1; p=0.025), ES: 0.50), all increases equivalent to small effects. Overall, these results demonstrate a sharp increase in symptom burden at illness onset, followed by gradual improvement, with residual symptoms persisting for several weeks.

### Subgroup characteristics and symptom trajectories

Patterns of symptom burden duration were broadly similar across age and risk groups, although high-risk participants had slightly shorter durations of moderate-to-severe and severe symptoms. In both younger (<50 years) and older (≥ 50 years) adults, durations for overall burden (composite score, mild-to-severe, and moderate-to-severe symptoms) were comparable to the overall cohort, while severe symptom durations were shorter. See Tables S3-S10 for detailed subgroup findings.

## DISCUSSION

This prospective outpatient study found substantial acute symptom burden at study enrollment (Day 1; corresponding to a median of 4 days since symptom onset, with variability across participants) followed by gradual improvements over the first month after diagnosis. Symptom prevalence and severity were high at enrollment, with widespread respiratory and systemic symptoms and large increases across all symptom-burden measures relative to pre-infection. Although symptom burden declined over time, composite symptom scores and counts of mild-to-severe, moderate-to-severe, and severe symptoms remained above baseline at Week 4 in the overall cohort. Patterns for composite scores and mild-to-severe, and moderate-to-severe symptoms were broadly similar across age- and risk-based strata, whereas severe symptoms showed earlier improvement (i.e. transitioned to moderate or mild severity), with timing varying by subgroup. By later time points, the absolute number of severe symptoms was low, but milder symptoms were present in a sizable proportion of individuals at Week 4, underscoring the ongoing clinical burden of COVID-19 in outpatient populations. These findings suggest that the time course of recovery was not uniform across subgroups, which could be driven by differences in vaccination status, antiviral treatment, and underlying risk profiles.

The findings on symptom duration align with prior research. In a retrospective cohort of 294 outpatients, the median symptom duration was 15 days, with over 25% symptomatic beyond three weeks [22]. Reported symptoms closely mirrored those among nonhospitalized patients in Colorado, where fatigue, cough, and myalgias were predominant early in illness [23]. However, that study was limited to a single US state and relied on retrospective telephone interviews conducted 10-52 days after symptom onset, requiring full illness recall. In contrast, the current study prospectively enrolled recently symptomatic, test-seeking adults within 0-4 days of symptom onset and captured symptoms in real time through surveys, minimizing recall-bias and enabling more granular characterization of symptom trajectories. Persistent symptom burden at one month aligns with growing evidence that some adults experience ongoing respiratory and fatigue-related symptoms beyond the acute phase [24,25].

Prior evidence of prolonged symptoms support examining changes in burden across cohorts. Symptom burden in the current cohort was lower than earlier COVID-19 cohorts from the same US retail pharmacy chain [11,13]. In the earliest cohort (January 31 to April 30, 2022) during circulation of Omicron BA.1.1, BA.2, and BA.2.12.1, the mean number of symptoms at Week 4 was 3.1 [13]. In a subsequent cohort (March 2 and May 18, 2023), during XBB 1.5 predominance, mean reported symptoms counts were 2.6 at Day 7, 1.9 at Day 14, and 0.9 at Week 4 [11]. The present cohort (October 24, 2024 and August 29, 2025) during JN.1 and XEC predominance showed correspondingly lower values of 1.9, 1.1, and 0.6. Together these data suggest progressively declining symptom burden from 2022 to 2024 consistent with lower severity associated with the JN.1 and XEC variants [26,27]. However, these comparisons are exploratory; future work should formally evaluate differences using harmonized symptom measures and accounting for SVI, population differences, and changes in vaccination, antiviral treatment, testing, and care-seeking.

In this study, adults ≥50 years experienced the longest persistence of severe symptoms across age groups. This aligns with evidence that older individuals have greater risk of severe illness and delayed recovery from respiratory infections such as influenza and COVID-19 [5,28]. A US-based registry study found older age, and comorbidities were associated with slower symptom recovery compared to younger ages and no comorbidities [9]. Persistent symptom burden in older adults has also been documented in studies with follow-up 3 to 9 months after COVID-19 illness onset (median ∼ 6 months) [29]. These age-related differences may reflect immunosenescence, reduced viral clearance, prolonged inflammation and higher comorbidity burden among participants ≥50 years (mean (SD) 1.2 (1.2) vs 0.9 (1.0) among those <50 years, p=0.002) (Table S5) that may delay recovery [30,31].

Notably, high-risk individuals experienced more rapid return to baseline of severe symptoms (by Day 4). This may reflect higher vaccination (19.9% vs 13.5%) and antiviral treatment (59.6% vs 44.5%) rates compared with those not at high risk (Table 1 and Table S5). Earlier antiviral treatment has shown to shorten symptom duration and reduce progression to severe disease [32], although, the timing of treatment initiation relative to symptom onset was not assessed. Future studies should evaluate whether earlier antiviral initiation or clinical engagement contributes to faster symptom resolution in high-risk populations [33,34].

Several limitations should be considered. This study relied on a convenience sample of outpatients who were reachable by email and willing to complete internet-based surveys, potentially limiting representativeness, particularly among individuals with limited digital access. The study population was predominantly female, White, employed, and resided primarily in the Southern United States, with relatively low social vulnerability, which may limit generalizability to more diverse or disadvantaged populations. Compared with adults with a positive COVID-19 test who were contacted by email but not included in the final analytic sample, primarily due to non-engagement, fewer were excluded for ineligibility, lack of enrollment or consent, or incomplete baseline data (Figure 1). Included participants were more likely to be female, White, and commercially insured, further limiting generalizability (Table S11). Recruitment through a retail pharmacy also underrepresent individuals seeking care through other healthcare channels.

Symptoms and symptom outcomes were self-reported and therefore subject to potential misclassification and recall bias, especially for pre-infection symptom reporting. While the mixed-effects model for repeated measures accounts for within-subject correlation over time, symptom severity ratings remain inherently subjective. and all symptoms were weighted equally despite differing clinical impact. The study period overlapped with circulation of other respiratory viruses, including influenza, which may have influenced symptom patterns and limits generalizability beyond similar seasonal contexts. Participants with more severe illness requiring hospitalization were likely underrepresented, potentially leading to underestimation of symptom severity and duration. Seventeen participants reported they did not feel well enough to respond, which may contribute to attrition bias (Table S12). Early symptom dynamics may be underrepresented as median enrollment occurred 4 days after symptom onset. Surveys were only available in English. Age and risk status were correlated, and subgroup analyses were not jointly stratified. Therefore, estimates may reflect underlying age or comorbidity differences and should be interpreted descriptively. Missing data were not imputed, and statistical testing was not adjusted for multiple comparisons, increasing the potential for Type I error. Participant compensation may have influenced survey completion or response behavior.

Despite these limitations, this study has important strengths. Participants were enrolled within 48 hours of testing positive, which could potentially minimize recall bias, and allow for more accurate characterization of early symptom severity and progression. The collection of pre-infection symptom data enabled direct estimation and quantification of symptom changes attributable to SARS-CoV-2 infection from baseline rather than enrollment. This study leveraged repeated, structured, graded symptom severity assessments in a large, test-confirmed COVID-19 outpatient cohort, enabling nuanced and clinically relevant characterization of COVID-19 symptom burden in non-hospitalized adults by age and risk status.

## CONCLUSION

In this prospective outpatient cohort, COVID-19 was associated with a pronounced acute symptom burden followed by incomplete recovery over the first month after diagnosis. Though some subgroups such as high-risk adults and adults younger than 50 years showed earlier improvement in severe symptoms. Overall symptom burden remained elevated through Week 4. Persistent symptom burden represents a clinically relevant component of acute morbidity and has implications for daily functioning, productivity, and patient counseling. Continued attention to lingering symptoms is warranted to inform outpatient care, recovery expectations, and supportive management strategies.

## Acknowledgements

LLL, RB and XS are employees of CVS Health Corporation, which received funding from Pfizer in connection with the development of this manuscript. Medical writing support was provided by Genevieve Meier at Aesara and Alexzandria Nicole Weikle at CVS Health and was funded by Pfizer.

## Data availability statement

Aggregated data that support the findings of this study are available upon reasonable request from the corresponding author Alon Yehoshua, subject to review. These data are not publicly available because they contain information that could compromise research participant privacy/consent.

## Ethics approval

The study was conducted according to the guidelines of the Declaration of Helsinki. The protocol, amendments and informed consent documents were reviewed and approved by the Sterling Institutional Review Board (Sterling IRB), an independent ethics committee based in Atlanta, Georgia, USA (Protocol #C4591034). The study was conducted in accordance with legal and regulatory requirements, as well as with scientific purpose, value, and rigor, and followed generally accepted research practices described in Good Practices for Outcomes Research issued by the Professional Society for Health Economics and Outcomes Research [35] and Good Pharmacoepidemiology Practice [36].

## Consent

Participation in the study was voluntary and anonymous. Consent was obtained from all subjects involved in the study electronically via the study’s E-Consent platform. Participants were informed of their right to refuse or withdraw from the study at any time. Participants were compensated for their time.

## Author contributions

All named authors meet the International Committee of Medical Journal Editors (ICMJE) criteria for authorship for this article.

Full list: A.Y., L.L.L., T.H., J.C.C., M.B.G., V.L.W., S.M.C.L., R.B., M.D.F., X.S.

Conceptualization: T.H., L.L.L., A.Y., J.C.C., V.L.W., M.D.F., X.S.

Methodology: T.H., L.L.L., A.Y., J.C.C., V.L.W., R.B., M.D.F., X.S.

Investigation: T.H., L.L.L., A.Y., J.C.C., V.L.W., M.D.F., X.S.

Resource: T.H., L.L.L., A.Y., M.B.G., X.S.

Data curation: X.S.

Formal analysis: T.H., A.Y., J.C.C., X.S.

Writing—original draft: R.B., X.S.

Writing—Review and editing: T.H., L.L.L., A.Y., J.C.C., M.B.G., V.L.W., R.B., M.D.F., X.S.

Supervision: T.H., A.Y., M.D.F.

Project Management: T.H., A.Y., M.B.G.

Funding Acquisition: T.H., A.Y., M.D.F.

All authors have read and agreed to the published version of the manuscript.

