## Supplemental material for "Quantifying the symptom burden of COVID-19: pre-infection through 1 month"

### Supporting Information

#### Supplementary Methods

##### CDC-identified conditions used to determine COVID-19 high-risk status

**Complete list of health conditions assessed during first survey**

During the first survey on Day 1 (enrollment day), participants were asked whether they received a diagnosis from a healthcare provider for any of the following comorbidities listed in the table below or are currently pregnant.

List of underlying health conditions associated with increased risk for COVID-19 complications:

- Any current/active cancers or malignancies (other than skin cancer)
- Cerebrovascular disease (such as stroke, transient ischemic attack [TIA])
- Chronic kidney disease (reduced kidney function or on dialysis)
- Chronic lung conditions (such as asthma, chronic obstructive pulmonary disease [COPD, emphysema], bronchiectasis, cystic fibrosis, pulmonary embolism [blood clots in lungs], pulmonary fibrosis, interstitial lung disease [ILD])
- Chronic liver disease (such as cirrhosis, fatty liver, alcoholic liver disease, autoimmune hepatitis)
- Endocrine disorders (such as diabetes [type 1 or type 2])
- Heart conditions (such as heart attack, heart failure, coronary artery disease, bypass surgery/stent, cardiomyopathy)
- Mental health conditions (such as mood disorders, depression, schizophrenia)
- Obesity: BMI>30
- Weakened immune system/ immunocompromised (currently on chemotherapy, chronic corticosteroids [such as prednisone], or anti-rejection medication due to an organ transplant; genetic immune disorders; other immune disorders, such as HIV or primary immunodeficiencies)
- Current or former smoker
- Active tuberculosis
- Pregnant

COVID-19=coronavirus disease 2019; COPD=chronic obstructive lung disease; BMI=body mass index; HIV=human immunodeficiency virus; ILD= interstitial lung disease; TIA=transient ischemic attack.

Supplementary Results Description

Subgroup characteristics and symptom trajectories

*High-risk participants*

A total of 379 participants (62.3%) met the COVID-19 high-risk criteria (mean (SD) age 48.5 (14.9) years; 77.8% female (Table 1). On Day 1, stuffy or runny nose (97.4%) and cough (94.5%) were the most common symptoms and remained most prevalent symptom through Week 4 (Table S3,).

Relative to the pre-infection period, symptom burden in this cohort increased substantially on Day 1, reflecting large effect sizes across all symptom measures (all p<0.001). The mean composite symptom score increased by 13.7 points (SE:0.4; ES: 2.12), mild-to-severe symptoms by 7.1 (SE: 0.2; ES: 2.50), moderate-to-severe symptoms by 5.0 (SE: 0.2; ES: 1.73), and severe symptoms by 1.7 (SE: 0.1; ES: 0.82) (Table S4).

Following this acute peak, symptom burden declined over time. The composite symptom score and the number of mild-to-severe and moderate-to-severe symptoms remained significantly elevated relative to pre-infection levels through Day 14 (all p<0.001, and all small ES). However, severe symptoms returned to baseline more rapidly, remaining significantly elevated only through Day 3 (p=0.020) (small ES of 0.24). Overall, symptom trajectories in the high-risk subgroup returned to baseline earlier than patterns observed in the total population.

*Adults ≥ 50 years*

Among the analytic sample, 244 (40.1%) participants were aged ≥50 years (Table S5). Within this subgroup, stuffy or runny nose and cough were the most frequently reported mild-to-severe symptoms at all timepoints (Table S6). On Day 1, symptom burden increased markedly relative to the pre-infection period (Table S7). The mean composite symptom score increased by 13.6 points (SE: 0.4; ES: 2.14), with corresponding increases of 7.5 (SE: 0.2; ES: 2.74) mild-to-severe, 4.8 (SE: 0.2; ES: 1.63) moderate-to-severe, and 1.6 (SE: 0.1; ES: 0.85) severe symptoms (all p<0.001). All within-cohort ES on Day 1 represent large effects.

Symptom burden declined progressively after Day 1. The composite symptom score, the number of mild-to-severe symptoms and moderate-to-severe symptoms remained significantly elevated through Week 4, with mean differences of 1.0 (SE: 0.2; ES: 0.34), 0.8 (SE: 0.2; ES: 0.34)), and 0.2 (SE: 0.1); ES: 0.20) (all p≤0.011). The number of severe symptoms remained elevated through Day 14 (mean difference: 0.2; SE: 0.1; p=0.032; ES: 0.48). These changes corresponded to small effect sizes.

*Adults <50 years*

The subgroup of adults aged <50 years included 364 (59.9%) participants (Table S5). As observed in other subgroups, mild-to-severe stuffy or runny nose and cough were the most prevalent symptoms (Supplementary Table 8).

On Day 1, symptom burden increased substantially relative to the pre-infection period. The mean composite symptom score increased by 14.7 points (SE: 0.3), with increases of 7.7 (SE: 0.1) mild-to-severe symptoms, 5.3 (SE: 0.1) moderate-to-severe symptoms, and 2.0 (SE: 0.1) severe symptoms (all p<0.001) (Table S7). Day 1 changes indicate large effects, evidenced by within-cohort effect sizes of 2.31 for the composite symptom score, 2.73 for mild-to-moderate symptoms, 1.92 for moderate-to-severe symptoms, and 1.00 for severe symptoms.

Symptom burden declined steadily after Day 1. The composite symptom score remained significantly elevated through Week 4 by 0.6 points (SE: 0.2; p<0.001), the number of mild-to-severe symptoms by 0.4 points (SE: 0.1; p=0.003), and the number of moderate-to-severe symptoms remained elevated through Week 4 by 0.2 (SE: 0.05; p<0.001). In contrast, severe symptoms returned to baseline more rapidly, remaining elevated only through Day 7 (mean difference: 0.2; SE: 0.1; p=0.024; ES: 0.35). Both changes correspond to small effects.

Patterns of symptom burden duration were broadly similar across age and risk groups, however, high-risk participants experienced slightly shorter moderate-to-severe and severe symptoms. Among adults younger than 50 years, the duration of overall burden as measured by the composite symptom score and counts of mild-to-severe, and moderate-to-severe symptoms was comparable to that of the overall cohort, while durations for severe symptoms were shorter. Similarly, among adults older than 50 years, the duration of overall burden as measured by the composite symptom score and counts of mild-to-severe, and moderate-to-severe symptoms was comparable to that of the overall cohort, while durations for severe symptoms were shorter. For all outcomes, observed mean scores and mean changes in the total population and across subgroups are presented in Tables S9 and S10.

Supplementary Results Tables

**Table S1**. Mean estimates for mixed-models for repeated measures for symptom outcomes (total population)

|  | **Composite acute symptom score** | | **Number of acute symptoms** | | **Number of moderate/ severe symptoms** | | **Number of severe symptoms** | |
| --- | --- | --- | --- | --- | --- | --- | --- | --- |
|  | **LSE (SE)** | **P-value** | **LSE (SE)** | **P-value** | **LSE (SE)** | **P-value** | **LSE (SE)** | **P-value** |
| **Intercept** | 19.90 (0.72) | <0.001 | 9.63 (0.39) | <0.001 | 7.01 (0.25) | <0.001 | 3.45 (0.21) | <0.001 |
| **Baseline score** | 0.54 (0.06) | <0.001 | 0.38 (0.04) | <0.001 | 0.73 (0.09) | <0.001 | 0.99 (0.26) | <0.001 |
| **Time** |  |  |  |  |  |  |  |  |
| Worst moment | Reference |  | Reference |  | Reference |  | Reference |  |
| Day 1 | -3.40 (0.37) | <0.001 | -0.63 (0.13) | <0.001 | -1.46 (0.18) | <0.001 | -1.40 (0.11) | <0.001 |
| Day 2 | -10.67 (0.43) | <0.001 | -3.35 (0.19) | <0.001 | -4.60 (0.20) | <0.001 | -2.79 (0.12) | <0.001 |
| Day 3 | -12.63 (0.43) | <0.001 | -4.58 (0.20) | <0.001 | -5.27 (0.20) | <0.001 | -2.86 (0.12) | <0.001 |
| Day 4 | -14.17 (0.43) | <0.001 | -5.38 (0.20) | <0.001 | -5.85 (0.19) | <0.001 | -3.06 (0.12) | <0.001 |
| Day 5 | -15.29 (0.43) | <0.001 | -6.17 (0.20) | <0.001 | -6.11 (0.19) | <0.001 | -3.13 (0.12) | <0.001 |
| Day 6 | -15.68 (0.43) | <0.001 | -6.52 (0.21) | <0.001 | -6.20 (0.19) | <0.001 | -3.12 (0.12) | <0.001 |
| Day 7 | -16.12 (0.44) | <0.001 | -6.90 (0.21) | <0.001 | -6.26 (0.19) | <0.001 | -3.10 (0.12) | <0.001 |
| Day 10 | -16.66 (0.45) | <0.001 | -7.28 (0.21) | <0.001 | -6.38 (0.19) | <0.001 | -3.16 (0.12) | <0.001 |
| Day 14 | -17.07 (0.45) | <0.001 | -7.71 (0.21) | <0.001 | -6.39 (0.19) | <0.001 | -3.11 (0.12) | <0.001 |
| Week 4 | -17.61 (0.45) | <0.001 | -8.19 (0.21) | <0.001 | -6.47 (0.18) | <0.001 | -3.13 (0.12) | <0.001 |
| **Clinical high-risk^a^ status** | -0.18 (1.68) | 0.916 | 0.11 (1.06) | 0.915 | 0.04 (0.52) | 0.937 | -0.11 (0.56) | 0.841 |
| **Clinical high-risk^a^ status by time** |  |  |  |  |  |  |  |  |
| Worst moment | Reference |  | Reference |  | Reference |  | Reference |  |
| Day 1 | -0.27 (0.44) | 0.537 | -0.10 (0.16) | 0.517 | -0.03 (0.21) | 0.902 | -0.07 (0.13) | 0.610 |
| Day 2 | 0.21 (0.52) | 0.684 | 0.20 (0.23) | 0.381 | 0.14 (0.24) | 0.554 | -0.08 (0.14) | 0.588 |
| Day 3 | 0.10 (0.51) | 0.839 | 0.47 (0.24) | 0.054 | 0.01 (0.23) | 0.965 | -0.34 (0.14) | 0.015 |
| Day 4 | 0.08 (0.51) | 0.870 | 0.40 (0.24) | 0.101 | 0.03 (0.23) | 0.905 | -0.34 (0.14) | 0.016 |
| Day 5 | -0.08 (0.51) | 0.875 | 0.34 (0.23) | 0.152 | -0.09 (0.23) | 0.700 | -0.35 (0.14) | 0.016 |
| Day 6 | -0.04 (0.52) | 0.943 | 0.38 (0.25) | 0.137 | -0.05 (0.22) | 0.836 | -0.39 (0.14) | 0.006 |
| Day 7 | 0.00 (0.53) | 0.994 | 0.45 (0.25) | 0.067 | -0.06 (0.23) | 0.791 | -0.42 (0.14) | 0.003 |
| Day 10 | -0.21 (0.54) | 0.705 | 0.34 (0.25) | 0.178 | -0.18 (0.23) | 0.435 | -0.46 (0.15) | 0.002 |
| Day 14 | -0.46 (0.55) | 0.401 | 0.18 (0.25) | 0.482 | -0.26 (0.23) | 0.264 | -0.51 (0.15) | 0.001 |
| Week 4 | -0.32 (0.54) | 0.556 | 0.36 (0.26) | 0.163 | -0.25 (0.22) | 0.266 | -0.52 (0.15) | 0.001 |
| **Age ≥50 years** | -0.96 (0.56) | 0.090 | -0.16 (0.20) | 0.424 | -0.39 (0.24) | 0.101 | -0.41 (0.12) | 0.001 |
| **Age ≥50 years by time** |  |  |  |  |  |  |  |  |
| Worst moment | Reference |  | Reference |  | Reference |  | Reference |  |
| Day 1 | 0.88 (0.47) | 0.062 | 0.30 (0.17) | 0.079 | 0.34 (0.22) | 0.135 | 0.27 (0.14) | 0.053 |
| Day 2 | 1.62 (0.55) | 0.003 | 0.65 (0.24) | 0.009 | 0.55 (0.25) | 0.029 | 0.43 (0.15) | 0.003 |
| Day 3 | 1.58 (0.54) | 0.004 | 0.67 (0.25) | 0.008 | 0.48 (0.25) | 0.051 | 0.42 (0.15) | 0.004 |
| Day 4 | 1.41 (0.54) | 0.009 | 0.51 (0.25) | 0.044 | 0.44 (0.24) | 0.069 | 0.51 (0.15) | 0.001 |
| Day 5 | 1.76 (0.54) | 0.001 | 0.84 (0.25) | 0.001 | 0.42 (0.24) | 0.084 | 0.53 (0.15) | <0.001 |
| Day 6 | 1.92 (0.55) | 0.001 | 0.92 (0.27) | 0.001 | 0.52 (0.24) | 0.027 | 0.50 (0.15) | 0.001 |
| Day 7 | 1.36 (0.56) | 0.015 | 0.58 (0.26) | 0.025 | 0.37 (0.24) | 0.121 | 0.43 (0.15) | 0.004 |
| Day 10 | 1.89 (0.57) | 0.001 | 0.79 (0.27) | 0.003 | 0.67 (0.24) | 0.006 | 0.50 (0.15) | 0.001 |
| Day 14 | 1.74 (0.57) | 0.003 | 0.82 (0.26) | 0.002 | 0.50 (0.24) | 0.040 | 0.48 (0.15) | 0.002 |
| Week 4 | 1.61 (0.57) | 0.005 | 0.72 (0.27) | 0.007 | 0.44 (0.23) | 0.060 | 0.47 (0.16) | 0.002 |
| **Self-reported sex** |  |  |  |  |  |  |  |  |
| Female | Reference |  | Reference |  | Reference |  | Reference |  |
| Male | -0.30 (0.27) | 0.271 | -0.24 (0.17) | 0.147 | -0.06 (0.08) | 0.455 | -0.13 (0.08) | 0.112 |
| Other | 4.06 (3.21) | 0.206 | 0.90 (1.79) | 0.617 | 0.79 (1.15) | 0.495 | -0.26 (0.81) | 0.752 |
| **Race/Ethnicity** |  |  |  |  |  |  |  |  |
| White | Reference |  | Reference |  | Reference |  | Reference |  |
| Black or African American | 0.58 (0.42) | 0.164 | -0.34 (0.26) | 0.191 | 0.01 (0.13) | 0.915 | 0.10 (0.13) | 0.464 |
| Hispanic or Latino | -0.17 (0.33) | 0.608 | -0.35 (0.21) | 0.091 | -0.03 (0.10) | 0.789 | -0.04 (0.11) | 0.731 |
| Asian | -0.53 (0.55) | 0.336 | -0.96 (0.34) | 0.005 | -0.10 (0.16) | 0.525 | -0.12 (0.17) | 0.490 |
| Other^b^ | -0.17 (0.61) | 0.783 | 0.06 (0.38) | 0.872 | -0.14 (0.18) | 0.448 | -0.03 (0.19) | 0.891 |
| **Social Vulnerability Index (SVI)^c^** |  |  |  |  |  |  |  |  |
| <0.25 | Reference |  | Reference |  | Reference |  | Reference |  |
| ≥0.25 and <0.5 | 0.17 (0.26) | 0.509 | 0.11 (0.16) | 0.515 | 0.07 (0.08) | 0.382 | 0.05 (0.08) | 0.552 |
| ≥0.5 and <0.75 | 0.02 (0.32) | 0.942 | 0.39 (0.20) | 0.055 | -0.05 (0.10) | 0.609 | 0.02 (0.10) | 0.849 |
| ≥0.75 | 1.05 (0.53) | 0.048 | 0.63 (0.33) | 0.058 | 0.38 (0.16) | 0.018 | 0.20 (0.17) | 0.251 |
| **US region** |  |  |  |  |  |  |  |  |
| Midwest | Reference |  | Reference |  | Reference |  | Reference |  |
| Northeast | 0.37 (0.38) | 0.332 | 0.26 (0.24) | 0.283 | 0.00 (0.11) | 0.999 | 0.05 (0.12) | 0.681 |
| South | 0.29 (0.34) | 0.394 | 0.34 (0.21) | 0.111 | 0.06 (0.10) | 0.583 | 0.02 (0.11) | 0.841 |
| West | 0.79 (0.55) | 0.153 | 0.34 (0.35) | 0.328 | 0.11 (0.17) | 0.520 | -0.14 (0.18) | 0.448 |
| Other/Unknown | -1.95 (3.25) | 0.549 | -0.63 (1.78) | 0.724 | -0.54 (1.17) | 0.643 | -0.27 (0.82) | 0.744 |
| **Number of comorbidities categories** |  |  |  |  |  |  |  |  |
| 0 | Reference |  | Reference |  | Reference |  | Reference | Reference |
| 1-2 | 0.79 (1.61) | 0.622 | -0.05 (1.06) | 0.964 | 0.30 (0.47) | 0.533 | 0.48 (0.55) | 0.79 (1.61) |
| 3+ | 2.58 (1.64) | 0.116 | 0.69 (1.07) | 0.522 | 0.82 (0.48) | 0.089 | 0.94 (0.56) | 2.58 (1.64) |
| **Insurance type** |  |  |  |  |  |  |  |  |
| Government | Reference |  | Reference |  | Reference |  | Reference | Reference |
| Commercial | -1.70 (0.49) | 0.001 | -0.82 (0.31) | 0.008 | -0.48 (0.15) | 0.001 | -0.24 (0.16) | -1.70 (0.49) |
| Self-pay | -1.67 (0.59) | 0.005 | -0.84 (0.37) | 0.024 | -0.61 (0.18) | 0.001 | -0.11 (0.19) | -1.67 (0.59) |

^a^Clinical high-risk criteria for COVID-19 (defined as adults aged ≥65 years or adults <65 years with ≥1 CDC-recognized underlying health condition associated with increased risk of COVID-19-related complications).

^b^American Indian or Alaska Native, Native Hawaiian or Other Pacific Islander, Multi-race, Prefer not to answer

^c^SVI is a score that ranges from 0 to 1. Higher values correspond to higher vulnerability.

**Table S2**. Prevalence of self-reported acute symptoms by severity from pre-infection to follow-up over a 1-month period among adults infected with COVID-19 in total population

| **Acute symptoms** | **Time of Survey, %** | | | | | | | | | | | |
| --- | --- | --- | --- | --- | --- | --- | --- | --- | --- | --- | --- | --- |
|  | **Prior to infection**  **(N=608)** | **Worst moment**  **(N=608)** | **Day 1**  **(N=608)** | **Day 2**  **(N=508)** | **Day 3 (N=516)** | **Day 4**  **(N=503)** | **Day 5**  **(N=474)** | **Day 6**  **(N=483)** | **Day 7**  **(N=472)** | **Day 10**  **(N=439)** | **Day 14**  **(N=432)** | **Week 4**  **(N=426)** |
| **Mild to severe, %** |  |  |  |  |  |  |  |  |  |  |  |  |
| stuffy or runny nose | 23.4 | 99.3 | 97.4 | 90.4 | 82.6 | 77.9 | 70.5 | 68.5 | 63.6 | 57.2 | 47.5 | 35.0 |
| cough | 8.4 | 94.6 | 94.1 | 87.4 | 79.7 | 73.2 | 66.2 | 60.0 | 56.4 | 52.2 | 44.0 | 30.3 |
| fatigue or tiredness | 27.8 | 94.7 | 93.6 | 83.5 | 76.4 | 68.2 | 59.7 | 53.8 | 48.5 | 44.0 | 36.8 | 28.9 |
| headache | 23.7 | 90.5 | 85.7 | 60.0 | 50.0 | 43.5 | 35.2 | 34.0 | 28.4 | 25.3 | 22.7 | 22.1 |
| body aches | 23.2 | 89.5 | 81.4 | 58.7 | 44.4 | 37.6 | 31.0 | 29.8 | 27.8 | 23.7 | 22.2 | 22.8 |
| feeling hot or feverish | 3.3 | 86.8 | 75.3 | 35.0 | 23.8 | 15.5 | 9.5 | 7.9 | 5.1 | 3.9 | 2.8 | 3.1 |
| chills or shivering | 2.6 | 80.1 | 64.0 | 23.0 | 13.2 | 8.4 | 5.5 | 5.4 | 4.5 | 4.1 | 2.6 | 2.4 |
| sore throat | 7.6 | 88.3 | 80.9 | 56.1 | 42.4 | 33.8 | 25.5 | 24.6 | 17.8 | 16.4 | 11.6 | 7.3 |
| difficulty breathing | 4.3 | 53.8 | 52.5 | 37.0 | 32.0 | 25.1 | 21.5 | 22.4 | 18.0 | 16.9 | 12.0 | 9.6 |
| nausea | 2.6 | 40.8 | 36.4 | 19.3 | 15.7 | 11.7 | 8.7 | 8.7 | 7.0 | 4.8 | 4.4 | 3.3 |
| vomit | 0.8 | 11.8 | 7.9 | 2.0 | 1.9 | 0.8 | 1.5 | 1.5 | 1.7 | 0.2 | 0.5 | 1.6 |
| diarrhea | 6.6 | 35.7 | 33.9 | 27.0 | 25.8 | 21.9 | 15.2 | 13.9 | 13.8 | 13.2 | 10.4 | 10.1 |
| change of smell | 1.3 | 35.7 | 33.1 | 25.6 | 22.5 | 19.7 | 16.5 | 13.5 | 11.7 | 8.0 | 5.3 | 3.1 |
| change of taste | 1.2 | 46.4 | 45.1 | 35.8 | 30.0 | 26.6 | 22.2 | 15.7 | 11.2 | 8.2 | 5.6 | 3.3 |
| **Moderate to severe, %** |  |  |  |  |  |  |  |  |  |  |  |  |
| stuffy or runny nose | 2.0 | 84.2 | 73.9 | 40.2 | 24.4 | 17.7 | 11.2 | 11.2 | 9.1 | 8.2 | 6.5 | 3.3 |
| cough | 0.5 | 72.0 | 61.5 | 36.8 | 25.2 | 18.5 | 12.0 | 13.3 | 10.6 | 10.9 | 7.2 | 4.2 |
| fatigue or tiredness | 2.5 | 81.9 | 74.2 | 41.1 | 28.3 | 21.5 | 16.2 | 13.5 | 11.0 | 10.5 | 6.3 | 7.0 |
| headache | 1.2 | 72.4 | 57.7 | 23.6 | 16.1 | 10.5 | 9.1 | 5.8 | 7.0 | 5.5 | 4.6 | 2.8 |
| body aches | 2.0 | 70.6 | 52.0 | 17.5 | 11.4 | 7.8 | 6.8 | 5.8 | 5.7 | 3.6 | 4.9 | 4.0 |
| feeling hot or feverish | 0.3 | 65.8 | 45.1 | 11.2 | 6.4 | 3.0 | 1.3 | 1.7 | 0.9 | 0.7 | 0.7 | 0.2 |
| chills or shivering | 0.2 | 63.0 | 38.2 | 6.3 | 2.7 | 2.4 | 0.8 | 1.0 | 1.3 | 0.5 | 0.5 | 0.2 |
| sore throat | 0.5 | 70.1 | 50.2 | 17.7 | 8.5 | 3.6 | 2.1 | 1.9 | 1.5 | 2.3 | 2.1 | 1.6 |
| difficulty breathing | 0.3 | 25.3 | 19.9 | 9.8 | 8.0 | 4.2 | 2.1 | 2.3 | 1.3 | 2.3 | 1.6 | 0.9 |
| nausea | 0.2 | 17.4 | 11.0 | 2.8 | 2.9 | 1.8 | 1.9 | 1.9 | 1.7 | 0.2 | 0.5 | 0.5 |
| vomit | 0.0 | 3.6 | 1.2 | 0.2 | 0.2 | 0.0 | 0.0 | 0.4 | 0.4 | 0.0 | 0.0 | 0.0 |
| diarrhea | 0.5 | 14.0 | 11.5 | 5.1 | 5.4 | 3.0 | 3.2 | 2.5 | 1.9 | 2.7 | 1.4 | 0.7 |
| change of smell | 0.7 | 12.7 | 11.4 | 10.6 | 9.7 | 7.8 | 5.5 | 5.0 | 4.7 | 2.5 | 2.1 | 1.4 |
| change of taste | 0.2 | 9.9 | 8.1 | 8.5 | 7.0 | 5.8 | 4.4 | 2.9 | 2.8 | 2.1 | 1.6 | 1.2 |
| **Severe, %** |  |  |  |  |  |  |  |  |  |  |  |  |
| stuffy or runny nose | 0.2 | 43.1 | 25.5 | 9.3 | 4.8 | 2.8 | 1.5 | 0.8 | 0.9 | 1.1 | 0.7 | 0.2 |
| cough | 0.0 | 29.0 | 18.1 | 7.1 | 4.3 | 2.6 | 1.7 | 1.7 | 1.1 | 0.7 | 0.7 | 1.2 |
| fatigue or tiredness | 0.2 | 44.6 | 32.7 | 9.3 | 4.7 | 2.4 | 1.9 | 1.2 | 0.4 | 0.7 | 1.4 | 0.7 |
| headache | 0.0 | 37.7 | 20.9 | 5.9 | 2.1 | 1.6 | 0.8 | 0.8 | 0.9 | 0.9 | 1.2 | 0.7 |
| body aches | 0.0 | 34.4 | 17.1 | 2.4 | 1.4 | 0.6 | 0.4 | 0.4 | 0.0 | 0.5 | 0.9 | 0.5 |
| feeling hot or feverish | 0.0 | 26.6 | 12.8 | 0.8 | 0.6 | 0.0 | 0.2 | 0.4 | 0.2 | 0.2 | 0.0 | 0.0 |
| chills or shivering | 0.0 | 26.8 | 11.0 | 1.2 | 0.2 | 0.0 | 0.0 | 0.2 | 0.2 | 0.2 | 0.2 | 0.0 |
| sore throat | 0.0 | 34.1 | 18.3 | 3.9 | 1.2 | 0.6 | 0.2 | 0.2 | 0.0 | 0.0 | 0.2 | 0.5 |
| difficulty breathing | 0.0 | 5.6 | 2.3 | 0.6 | 0.6 | 0.4 | 0.2 | 0.0 | 0.0 | 0.0 | 0.0 | 0.0 |
| nausea | 0.0 | 4.1 | 2.0 | 0.2 | 0.8 | 0.2 | 0.4 | 0.8 | 0.4 | 0.0 | 0.0 | 0.0 |
| vomit | 0.0 | 1.0 | 0.5 | 0.0 | 0.0 | 0.0 | 0.0 | 0.4 | 0.0 | 0.0 | 0.0 | 0.0 |
| diarrhea | 0.2 | 4.0 | 2.6 | 1.2 | 2.1 | 0.8 | 0.4 | 1.0 | 0.6 | 0.2 | 0.5 | 0.2 |
| change of smell | 0.7 | 12.7 | 11.4 | 10.6 | 9.7 | 7.7 | 5.5 | 5.0 | 4.7 | 2.5 | 2.1 | 1.4 |
| change of taste | 0.2 | 9.9 | 8.1 | 8.4 | 7.0 | 5.7 | 4.4 | 2.9 | 2.8 | 2.1 | 1.6 | 1.2 |

**Supplementary Table S3**. Prevalence of self-reported acute symptoms by severity from pre-infection to follow-up over a 1-month period among adults at high risk for severe complications of COVID-19*

| **Acute symptoms** | **Time of Survey. %** | | | | | | | | | | | |
| --- | --- | --- | --- | --- | --- | --- | --- | --- | --- | --- | --- | --- |
|  | **Prior to infection**  **(N=379)** | **Worst moment**  **(N=379)** | **Day 1**  **(N=379)** | **Day 2**  **(N=321)** | **Day 3 (N=327)** | **Day 4**  **(N=319)** | **Day 5**  **(N=308)** | **Day 6**  **(N=310)** | **Day 7**  **(N=303)** | **Day 10**  **(N=285)** | **Day 14**  **(N=283)** | **Week 4**  **(N=278)** |
| **Mild to severe, %** |  |  |  |  |  |  |  |  |  |  |  |  |
| stuffy or runny nose | 25.6 | 99.5 | 97.4 | 90.0 | 83.5 | 76.8 | 70.5 | 70.3 | 66.0 | 58.3 | 49.8 | 25.6 |
| cough | 9.8 | 96.0 | 94.5 | 87.2 | 79.8 | 73.4 | 69.8 | 62.6 | 59.1 | 53.0 | 43.8 | 9.8 |
| fatigue or tiredness | 34.8 | 95.5 | 94.2 | 83.2 | 78.3 | 73.7 | 64.0 | 56.8 | 53.8 | 49.8 | 42.1 | 34.8 |
| headache | 28.8 | 91.8 | 87.1 | 62.9 | 53.5 | 45.8 | 37.7 | 36.8 | 33.0 | 29.5 | 26.5 | 28.8 |
| body aches | 29.6 | 89.5 | 81.8 | 62.3 | 49.2 | 43.0 | 36.4 | 33.9 | 33.7 | 30.2 | 27.6 | 29.6 |
| feeling hot or feverish | 4.5 | 86.8 | 73.6 | 37.4 | 26.3 | 18.2 | 12.3 | 9.7 | 5.9 | 4.6 | 3.5 | 4.5 |
| chills or shivering | 3.4 | 80.0 | 60.7 | 26.2 | 15.6 | 10.7 | 5.8 | 6.5 | 6.3 | 4.9 | 3.5 | 3.4 |
| sore throat | 6.9 | 86.0 | 78.9 | 55.1 | 43.1 | 35.1 | 25.3 | 24.8 | 19.1 | 16.8 | 11.0 | 6.9 |
| difficulty breathing | 5.8 | 54.1 | 53.6 | 38.9 | 35.5 | 27.0 | 24.0 | 25.5 | 20.8 | 19.3 | 13.4 | 5.8 |
| nausea | 3.7 | 43.0 | 38.8 | 19.3 | 17.4 | 12.5 | 9.4 | 8.4 | 7.6 | 4.6 | 4.2 | 3.7 |
| vomit | 0.8 | 14.3 | 9.2 | 2.2 | 2.8 | 0.9 | 2.0 | 1.9 | 2.0 | 0.0 | 0.7 | 0.8 |
| diarrhea | 8.2 | 38.0 | 36.4 | 29.6 | 28.1 | 24.1 | 15.9 | 16.1 | 15.2 | 14.0 | 10.3 | 8.2 |
| change of smell | 1.1 | 39.6 | 37.5 | 27.4 | 22.9 | 20.4 | 16.6 | 13.9 | 12.9 | 9.1 | 6.4 | 1.1 |
| change of taste | 1.1 | 47.8 | 46.7 | 37.4 | 31.8 | 28.5 | 22.7 | 15.8 | 12.9 | 9.8 | 6.7 | 1.1 |
| **Moderate to severe, %** |  |  |  |  |  |  |  |  |  |  |  |  |
| stuffy or runny nose | 2.37 | 85.49 | 74.67 | 44.24 | 25.99 | 19.75 | 10.71 | 12.58 | 11.22 | 10.18 | 7.77 | 4.32 |
| cough | 0.79 | 73.88 | 62.01 | 35.51 | 25.99 | 20.06 | 13.31 | 13.23 | 12.54 | 11.23 | 6.36 | 3.96 |
| fatigue or tiredness | 3.43 | 82.85 | 76.25 | 43.93 | 32.11 | 24.45 | 19.16 | 16.45 | 13.53 | 14.04 | 8.83 | 10.07 |
| headache | 1.32 | 73.61 | 58.05 | 25.23 | 16.21 | 11.91 | 10.06 | 7.10 | 8.58 | 7.37 | 5.65 | 3.24 |
| body aches | 2.11 | 72.30 | 53.03 | 19.31 | 13.15 | 9.72 | 8.44 | 7.10 | 6.93 | 4.56 | 6.36 | 5.40 |
| feeling hot or feverish | 0.53 | 63.85 | 43.54 | 13.08 | 5.50 | 3.45 | 1.95 | 2.26 | 1.32 | 0.70 | 0.71 | 0.36 |
| chills or shivering | 0.26 | 63.32 | 36.68 | 7.48 | 2.75 | 2.82 | 1.30 | 1.29 | 1.65 | 0.70 | 0.71 | 0.36 |
| sore throat | 0.53 | 68.34 | 46.70 | 17.13 | 10.09 | 4.08 | 2.27 | 2.58 | 1.98 | 2.46 | 2.12 | 1.44 |
| difficulty breathing | 0.26 | 27.44 | 22.43 | 9.97 | 9.17 | 5.02 | 2.92 | 3.23 | 1.65 | 2.46 | 2.12 | 1.08 |
| nausea | 0.26 | 18.47 | 10.82 | 2.80 | 3.06 | 2.51 | 2.92 | 2.26 | 2.31 | 0.35 | 0.71 | 0.72 |
| vomit | 0.00 | 4.22 | 1.58 | 0.31 | 0.31 | 0.00 | 0.00 | 0.65 | 0.33 | 0.00 | 0.00 | 0.00 |
| diarrhea | 0.53 | 15.30 | 12.93 | 5.30 | 5.50 | 3.13 | 3.90 | 2.58 | 2.64 | 3.16 | 1.77 | 1.08 |
| change of smell | 0.26 | 13.19 | 13.19 | 12.15 | 11.31 | 8.46 | 6.17 | 5.48 | 6.27 | 3.16 | 2.47 | 1.80 |
| change of taste | 0.00 | 10.03 | 9.76 | 10.28 | 7.03 | 5.64 | 4.87 | 2.90 | 3.63 | 2.46 | 1.77 | 1.44 |
| **Severe, %** |  |  |  |  |  |  |  |  |  |  |  |  |
| stuffy or runny nose | 0.26 | 44.33 | 26.91 | 9.97 | 4.89 | 3.13 | 1.62 | 0.97 | 0.99 | 1.40 | 0.35 | 0.36 |
| cough | 0.00 | 31.13 | 16.89 | 8.10 | 4.59 | 2.19 | 2.27 | 0.65 | 0.99 | 0.70 | 0.35 | 0.72 |
| fatigue or tiredness | 0.26 | 48.02 | 35.62 | 10.59 | 4.89 | 2.82 | 2.92 | 1.94 | 0.66 | 1.05 | 2.12 | 1.08 |
| headache | 0.00 | 40.11 | 20.84 | 7.48 | 2.45 | 2.19 | 1.30 | 0.97 | 1.32 | 1.40 | 1.06 | 0.72 |
| body aches | 0.00 | 35.36 | 17.68 | 3.43 | 1.53 | 0.94 | 0.32 | 0.65 | 0.00 | 0.70 | 1.41 | 0.72 |
| feeling hot or feverish | 0.00 | 25.86 | 13.46 | 0.93 | 0.31 | 0.00 | 0.32 | 0.65 | 0.33 | 0.35 | 0.00 | 0.00 |
| chills or shivering | 0.00 | 25.07 | 10.29 | 1.25 | 0.00 | 0.00 | 0.00 | 0.32 | 0.33 | 0.35 | 0.35 | 0.00 |
| sore throat | 0.00 | 31.40 | 16.62 | 4.67 | 1.22 | 0.94 | 0.32 | 0.32 | 0.00 | 0.00 | 0.35 | 0.36 |
| difficulty breathing | 0.00 | 5.54 | 2.11 | 0.62 | 0.31 | 0.31 | 0.32 | 0.00 | 0.00 | 0.00 | 0.00 | 0.00 |
| nausea | 0.00 | 4.22 | 2.37 | 0.31 | 0.92 | 0.31 | 0.65 | 1.29 | 0.66 | 0.00 | 0.00 | 0.00 |
| vomit | 0.00 | 1.32 | 0.53 | 0.00 | 0.00 | 0.00 | 0.00 | 0.65 | 0.00 | 0.00 | 0.00 | 0.00 |
| diarrhea | 0.26 | 4.75 | 3.17 | 1.56 | 2.14 | 1.25 | 0.65 | 0.97 | 0.66 | 0.35 | 0.71 | 0.36 |
| change of smell | 0.26 | 13.19 | 13.19 | 12.07 | 11.31 | 8.41 | 6.15 | 5.47 | 6.25 | 3.16 | 2.47 | 1.80 |
| change of taste | 0.00 | 10.03 | 9.76 | 10.22 | 7.03 | 5.61 | 4.85 | 2.89 | 3.62 | 2.46 | 1.77 | 1.44 |

*Defined as adults aged ≥65 years or adults <65 years with ≥1 CDC-recognized underlying health condition associated with increased risk of COVID-19-related complications

**Table S4**. Summary of least-square estimates^a^ of mean change in symptom measures from prior to infection in high-risk* group.

| **Time** | **Retention rate**  **n (%)** | **Composite acute symptom score** | | | **Number of mild to severe acute symptoms** | | | **Number of moderate to severe acute symptoms** | | | **Number of severe acute symptoms** | | |
| --- | --- | --- | --- | --- | --- | --- | --- | --- | --- | --- | --- | --- | --- |
|  |  | **LSE (SE)** | **P-value** | **ES^b^** | **LSE (SE)** | **P-value** | **ES^b^** | **LSE (SE)** | **P-value** | **ES^b^** | **LSE (SE)** | **P-value** | **ES^b^** |
| **High-risk population** | | | | | | | | | | | | | |
| Prior to infection^c^ | 379 (100.0) | 1.8 (2.0) |  |  | 1.6 (1.7) | . |  | 0.1 (0.5) | . |  | 0.0 (0.1) | . |  |
| Worst Moment | 379 (100.0) | 17.2 (0.4) | **<0.001** | 2.73 | 7.9 (0.2) | **<0.001** | 3.12 | 6.5 (0.1) | **<0.001** | 2.46 | **3.0 (0.1)** | **<0.001** | 1.15 |
| Day 1 | 379 (100.0) | 13.7 (0.4) | **<0.001** | 2.12 | 7.1 (0.2) | **<0.001** | 2.50 | 5.0 (0.2) | **<0.001** | 1.73 | **1.7 (0.1)** | **<0.001** | 0.82 |
| Day 2 | 321 (84.7) | 7.5 (0.3) | **<0.001** | 1.34 | 4.8 (0.2) | **<0.001** | 1.64 | 2.2 (0.1) | **<0.001** | 0.91 | **0.5 (0.1)** | **<0.001** | 0.40 |
| Day 3 | 327 (86.3) | 5.5 (0.3) | **<0.001** | 1.15 | 3.9 (0.2) | **<0.001** | 1.39 | 1.4 (0.1) | **<0.001** | 0.71 | **0.2 (0.1)** | **0.020** | 0.24 |
| Day 4 | 319 (84.2) | 4.2 (0.3) | **<0.001** | 1.03 | 3.2 (0.2) | **<0.001** | 1.18 | 1.0 (0.1) | **<0.001** | 0.59 | 0.1 (0.1) | 0.287 | 0.14 |
| Day 5 | 308 (81.3) | 3.1 (0.3) | **<0.001** | 0.83 | 2.4 (0.2) | **<0.001** | 0.96 | 0.7 (0.1) | **<0.001** | 0.44 | 0.03 (0.09) | 0.770 | 0.04 |
| Day 6 | 310 (81.8) | 2.8 (0.3) | **<0.001** | 0.73 | 2.1 (0.2) | **<0.001** | 0.81 | 0.6 (0.1) | **<0.001** | 0.42 | -0.02 (0.09) | 0.829 | -0.03 |
| Day 7 | 303 (79.9) | 2.2 (0.3) | **<0.001** | 0.60 | 1.7 (0.2) | **<0.001** | 0.69 | 0.5 (0.1) | **<0.001** | 0.36 | -0.05 (0.09) | 0.568 | -0.09 |
| Day 10 | 285 (75.2) | 1.8 (0.3) | **<0.001** | 0.54 | 1.4 (0.2) | **<0.001** | 0.58 | 0.4 (0.1) | **<0.001** | 0.34 | -0.1 (0.1) | 0.341 | -0.20 |
| Day 14 | 283 (74.7) | 1.1 (0.2) | **<0.001** | 0.36 | 0.8 (0.2) | **<0.001** | 0.37 | 0.2 (0.1) | **0.002** | 0.24 | -0.1 (0.1) | 0.279 | -0.24 |
| Week 4 | 278 (73.4) | 0.5 (0.2) | **0.024** | 0.19 | 0.4 (0.2) | **0.012** | 0.19 | 0.1 (0.1) | 0.054 | 0.16 | -0.1 (0.1) | 0.165 | -0.37 |

LSE=least-square estimate; SE=standard error; ES=effect size.

*Defined as adults aged ≥65 years or adults <65 years with ≥1 CDC-recognized underlying health condition associated with increased risk of COVID-19-related complications.

^a^ Least-square estimates based on mixed models for repeated measures, which include covariates of time, high-risk status, self-reported sex, age group (≥50 vs <50), race/ethnicity, US region, social vulnerability index category, baseline outcome score, insurance type, number of comorbidities, and interaction terms between time and high-risk status, as well as time and age group, with unstructured correlation matrix for repeated measures.

^b^ ES refers to within-cohort effect size, was calculated as the least square estimate of mean change scores divided by the observed standard deviation of change scores from prior to infection to follow-up.

^c^ Mean (standard deviation) of scores prior to infection reflect observed values and were not derived using the least squares model estimates.

Bolded values indicate P<0.05 which is considered statistically significant.

**Table S5. Baseline characteristics of adults not at high-risk for COVID, adults ≥50 years and <50 years**

|  | **Not at high-risk for COVID-19**  **(N=229)** | **Adults ≥50 years**  **(N=244)** | **Adults <50 years**  **(N=364)** |
| --- | --- | --- | --- |
| Age, years, mean (SD) | 41.5 (11.7) | 60.3 (7.5) | 36.1 (7.9) |
| Self-reported sex, n (%) |  |  |  |
| Female | 170 (74.2) | 180 (73.8) | 285 (78.3) |
| Race/Ethnicity, n (%) |  |  |  |
| White | 148 (64.6) | 192 (78.7) | 244 (67.0) |
| Black or African American | 15 (6.6) | 18 (7.4) | 30 (8.2) |
| Hispanic or Latino | 43 (18.8) | 19 (7.8) | 60 (16.5) |
| Asian | 14 (6.1) | 11 (4.5) | 14 (3.8) |
| Other | 9 (3.9) | 4 (1.6) | 16 (4.4) |
| US Region of residence, n (%) |  |  |  |
| Northeast | 31 (13.5) | 32 (13.1) | 41 (11.3) |
| South | 138 (60.3) | 140 (57.4) | 228 (62.6) |
| Midwest | 45 (19.7) | 57 (23.4) | 76 (20.9) |
| West | 14 (6.1) | 14 (5.7) | 19 (5.2) |
| Other\Unknown | 1 (0.4) | 1 (0.4) | 0 (0.0) |
| Employment status, n (%) |  |  |  |
| Yes | 193 (84.3) | 164 (67.2) | 312 (85.7) |
| Social Vulnerability Index ^a^, mean (SD) | 0.36 (0.20) | 0.35 (0.21) | 0.38 (0.21) |
| Social Vulnerability Index category^a^, n (%) |  |  |  |
| <0.25 | 85 (37.1) | 91 (37.3) | 114 (31.3) |
| ≥0.25 and <0.5 | 87 (38.0) | 97 (39.8) | 147 (40.4) |
| ≥0.5 and <0.75 | 48 (21.0) | 46 (18.9) | 83 (22.8) |
| ≥0.75 | 9 (3.9) | 10 (4.1) | 20 (5.5) |
| Antiviral use, n (%) | 102 (44.5) | 159 (65.2) | 169 (46.4) |
| Received 2024-2025 Pfizer-BioNTech vaccine, n (%) | 31 (13.5) | 58 (23.8) | 48 (13.2) |
| Pregnant, n (%) |  |  |  |
| No | 168 (73.4) | 127 (52.0) | 287 (78.8) |
| Yes | 0 (0.0) | 0 (0.0) | 5 (1.4) |
| Not applicable | 61 (26.6) | 117 (48.0) | 72 (19.8) |
| Underlying health conditions, n (%) |  |  |  |
| Any current/active cancers or malignancies^c^ | 0 (0.0) | 5 (2.0) | 2 (0.5) |
| Cerebrovascular disease | 0 (0.0) | 4 (1.6) | 3 (0.8) |
| Chronic kidney disease | 0 (0.0) | 7 (2.9) | 0 (0.0) |
| Chronic lung conditions | 0 (0.0) | 28 (11.5) | 37 (10.2) |
| Chronic liver disease | 0 (0.0) | 8 (3.3) | 4 (1.1) |
| Endocrine disorders | 0 (0.0) | 33 (13.5) | 18 (4.9) |
| Heart conditions | 0 (0.0) | 14 (5.7) | 8 (2.2) |
| Mental health conditions | 0 (0.0) | 39 (16.0) | 103 (28.3) |
| Obesity: BMI>30 | 0 (0.0) | 84 (34.4) | 99 (27.2) |
| Weakened immune system/ immunocompromised | 0 (0.0) | 11 (4.5) | 14 (3.8) |
| Smoker | 0 (0.0) | 53 (21.7) | 35 (9.6) |
| Active tuberculosis | 0 (0.0) | (0.0) | (0.0) |
| Number of comorbidities, mean (SD) | 0.0 (0.0) | 1.2 (1.2) | 0.9 (1.0) |
| ≥1 comorbidity, n (%) | 0 (0.0) | 153 (62.7) | 199 (54.7) |
| ≥1 comorbidity or pregnant, n (%) | 0 (0.0) | 153 (62.7) | 201 (55.2) |
| Time from symptom onset to enrollment, days, Median (IQR) | 4.0 (3~5) | 4.0 (3~5) | 4.0 (3~5) |
| Time from vaccination to enrollment, days, Median (IQR) | 1102.0 (553~1,324) | 749.5 (301.5~1,188.5) | 1087.5 (568.500~1,321.500) |

BMI=body mass index; IQR=Interquartile range; SD=standard deviation; US=United States.

^a^SVI is a score that ranges from 0 to 1. Higher values correspond to higher vulnerability.

^b^High-risk criteria for COVID-19 (defined as adults aged ≥65 years and adults <65 years with ≥1 CDC-recognized underlying health condition associated with increased risk of getting very sick from COVID-19).

^c^Any current/active cancers or malignancies (other than skin cancer)

**Table S6**. Prevalence of self-reported acute symptoms by severity from pre-infection to follow-up over a 1-month period among adults infected with COVID-19 aged ≥50 years

| **Acute symptoms** | **Time of Survey, %** | | | | | | | | | | | |
| --- | --- | --- | --- | --- | --- | --- | --- | --- | --- | --- | --- | --- |
|  | **Prior to infection**  **(N=244)** | **Worst moment**  **(N=244)** | **Day 1**  **(N=244)** | **Day 2**  **(N=201)** | **Day 3 (N=204)** | **Day 4**  **(N=197)** | **Day 5**  **(N=190)** | **Day 6**  **(N=190)** | **Day 7**  **(N=194)** | **Day 10**  **(N=178)** | **Day 14**  **(N=176)** | **Week 4**  **(N=170)** |
| **Mild to severe, %** |  |  |  |  |  |  |  |  |  |  |  |  |
| stuffy or runny nose | 22.5 | 99.6 | 97.1 | 90.6 | 83.3 | 75.1 | 70.5 | 73.2 | 68.6 | 59.6 | 54.0 | 37.1 |
| cough | 9.8 | 96.3 | 95.5 | 87.1 | 80.9 | 72.1 | 70.5 | 64.2 | 59.3 | 55.1 | 47.7 | 35.9 |
| fatigue or tiredness | 27.9 | 94.7 | 93.9 | 85.6 | 77.9 | 76.1 | 67.4 | 57.4 | 52.6 | 49.4 | 44.3 | 33.5 |
| headache | 18.0 | 88.5 | 83.2 | 57.2 | 48.0 | 43.2 | 34.7 | 35.3 | 29.9 | 29.8 | 27.3 | 22.4 |
| body aches | 29.1 | 87.3 | 80.7 | 62.7 | 48.5 | 43.7 | 32.6 | 32.6 | 30.4 | 28.1 | 27.8 | 32.9 |
| feeling hot or feverish | 3.3 | 83.2 | 73.0 | 36.3 | 26.5 | 17.3 | 11.1 | 9.0 | 4.6 | 5.1 | 2.8 | 2.4 |
| chills or shivering | 2.9 | 77.5 | 61.9 | 23.4 | 11.8 | 7.6 | 5.8 | 7.4 | 4.6 | 3.9 | 3.4 | 2.9 |
| sore throat | 6.6 | 83.6 | 76.6 | 56.7 | 36.8 | 31.5 | 24.2 | 25.3 | 16.0 | 20.2 | 13.6 | 7.7 |
| difficulty breathing | 5.7 | 47.5 | 47.1 | 34.3 | 32.4 | 25.9 | 25.3 | 28.4 | 19.6 | 20.2 | 13.1 | 12.4 |
| nausea | 2.9 | 37.7 | 35.3 | 19.9 | 18.6 | 9.6 | 9.5 | 7.4 | 7.7 | 6.2 | 5.7 | 4.7 |
| vomit | 0.0 | 12.3 | 7.8 | 1.0 | 2.5 | 1.0 | 1.6 | 2.1 | 2.1 | 0.0 | 0.0 | 1.8 |
| diarrhea | 5.3 | 34.0 | 33.2 | 28.4 | 24.0 | 22.3 | 13.2 | 12.6 | 12.9 | 14.6 | 9.1 | 10.6 |
| change of smell | 2.1 | 34.4 | 32.0 | 25.9 | 20.6 | 19.3 | 17.4 | 14.7 | 12.4 | 10.1 | 8.0 | 5.3 |
| change of taste | 1.2 | 45.9 | 48.4 | 38.3 | 34.8 | 30.5 | 27.9 | 20.0 | 14.4 | 11.2 | 8.5 | 5.9 |
| **Moderate to severe, %** |  |  |  |  |  |  |  |  |  |  |  |  |
| stuffy or runny nose | 2.5 | 82.8 | 73.4 | 39.8 | 23.5 | 14.7 | 11.6 | 11.1 | 8.8 | 10.1 | 7.4 | 4.1 |
| cough | 1.2 | 75.0 | 64.3 | 36.3 | 24.5 | 18.3 | 12.6 | 14.7 | 11.9 | 12.9 | 7.4 | 4.7 |
| fatigue or tiredness | 3.3 | 78.7 | 71.7 | 38.8 | 27.0 | 21.3 | 14.7 | 14.2 | 9.8 | 14.6 | 7.4 | 8.2 |
| headache | 1.2 | 69.3 | 51.2 | 20.9 | 13.2 | 10.2 | 8.4 | 5.3 | 5.2 | 5.6 | 3.4 | 1.8 |
| body aches | 3.3 | 66.0 | 49.6 | 19.4 | 11.3 | 8.6 | 5.3 | 3.7 | 4.1 | 4.5 | 5.1 | 4.7 |
| feeling hot or feverish | 0.4 | 59.0 | 39.3 | 12.4 | 5.9 | 4.1 | 0.5 | 0.5 | 0.5 | 0.0 | 0.6 | 0.0 |
| chills or shivering | 0.4 | 55.7 | 36.9 | 6.5 | 2.9 | 3.6 | 1.1 | 0.5 | 1.0 | 0.0 | 1.1 | 0.0 |
| sore throat | 0.4 | 63.1 | 44.3 | 15.9 | 7.4 | 4.6 | 2.6 | 2.6 | 2.1 | 2.3 | 2.3 | 1.2 |
| difficulty breathing | 0.4 | 23.8 | 17.2 | 9.0 | 6.9 | 4.6 | 2.6 | 3.2 | 1.6 | 4.5 | 1.7 | 0.6 |
| nausea | 0.0 | 16.8 | 11.9 | 4.0 | 3.4 | 1.0 | 1.6 | 1.6 | 1.6 | 0.6 | 0.6 | 0.6 |
| vomit | 0.0 | 4.9 | 1.6 | 0.0 | 0.5 | 0.0 | 0.0 | 1.1 | 0.0 | 0.0 | 0.0 | 0.0 |
| diarrhea | 0.0 | 13.9 | 9.4 | 5.5 | 4.9 | 2.5 | 2.6 | 1.6 | 1.0 | 2.3 | 1.7 | 0.6 |
| change of smell | 0.4 | 10.7 | 10.3 | 10.0 | 8.8 | 8.1 | 6.3 | 6.3 | 6.2 | 4.5 | 4.6 | 2.9 |
| change of taste | 0.0 | 10.3 | 9.4 | 9.0 | 7.4 | 6.1 | 4.2 | 3.7 | 3.6 | 3.9 | 3.4 | 2.4 |
| **Severe, %** |  |  |  |  |  |  |  |  |  |  |  |  |
| stuffy or runny nose | 0.00 | 39.75 | 23.77 | 8.96 | 3.43 | 3.55 | 2.11 | 1.05 | 1.03 | 2.25 | 1.14 | 0.00 |
| cough | 0.00 | 31.15 | 18.44 | 6.97 | 3.43 | 3.05 | 2.11 | 2.11 | 1.03 | 1.12 | 1.14 | 2.35 |
| fatigue or tiredness | 0.41 | 38.52 | 28.28 | 5.97 | 3.43 | 2.03 | 2.63 | 1.58 | 0.00 | 1.12 | 1.14 | 0.59 |
| headache | 0.00 | 35.66 | 18.03 | 4.98 | 2.45 | 1.02 | 1.58 | 0.53 | 0.00 | 0.56 | 0.00 | 0.00 |
| body aches | 0.00 | 30.33 | 12.70 | 1.49 | 0.49 | 0.00 | 0.00 | 0.00 | 0.00 | 0.00 | 0.57 | 1.18 |
| feeling hot or feverish | 0.00 | 16.80 | 9.43 | 0.50 | 0.00 | 0.00 | 0.00 | 0.00 | 0.00 | 0.00 | 0.00 | 0.00 |
| chills or shivering | 0.00 | 19.26 | 7.38 | 0.50 | 0.00 | 0.00 | 0.00 | 0.00 | 0.00 | 0.00 | 0.57 | 0.00 |
| sore throat | 0.00 | 31.97 | 15.57 | 5.47 | 0.98 | 1.02 | 0.53 | 0.00 | 0.00 | 0.00 | 0.00 | 0.00 |
| difficulty breathing | 0.00 | 3.28 | 1.23 | 1.00 | 0.98 | 0.51 | 0.53 | 0.00 | 0.00 | 0.00 | 0.00 | 0.00 |
| nausea | 0.00 | 3.28 | 2.46 | 0.50 | 0.49 | 0.51 | 0.53 | 1.58 | 0.00 | 0.00 | 0.00 | 0.00 |
| vomit | 0.00 | 0.82 | 0.82 | 0.00 | 0.00 | 0.00 | 0.00 | 1.05 | 0.00 | 0.00 | 0.00 | 0.00 |
| diarrhea | 0.00 | 2.87 | 2.05 | 1.00 | 1.96 | 1.02 | 0.53 | 0.53 | 0.52 | 0.56 | 0.57 | 0.59 |
| change of smell | 0.41 | 10.66 | 10.25 | 9.90 | 8.82 | 8.08 | 6.32 | 6.28 | 6.19 | 4.49 | 4.55 | 2.94 |
| change of taste | 0.00 | 10.25 | 9.43 | 8.91 | 7.35 | 6.06 | 4.21 | 3.66 | 3.61 | 3.93 | 3.41 | 2.35 |

**Table S7**. Summary of least-square estimates^a^ of mean change in symptom measures from prior to infection in adults ≥50 years and <50 years.

| **Time** | **Retention rate**  **n (%)** | **Composite symptom score** | | | **Number of mild to severe acute symptoms** | | | **Number of moderate to severe acute symptoms** | | | **Number of severe acute symptoms** | | |
| --- | --- | --- | --- | --- | --- | --- | --- | --- | --- | --- | --- | --- | --- |
|  |  | **LSE (SE)** | **P-value** | **ES^b^** | **LSE (SE)** | **P-value** | **ES^b^** | **LSE (SE)** | **P-value** | **ES^b^** | **LSE (SE)** | **P-value** | **ES^b^** |
| **Adults ≥50 years** | | | | | | | | | | | | | |
| Prior to infection^c^ | 244 (100.0) | 1.5 (1.9) |  |  | 1.4 (1.6) | . |  | 0.1 (0.5) | . |  | 0.0 (0.1) | . |  |
| Worst Moment | 244 (100.0) | 16.6 (0.4) | **<0.001** | 2.69 | **8.0 (0.2)** | 0.000 | 3.40 | **6.1 (0.2)** | 0.000 | 2.21 | **2.8 (0.1)** | 0.000 | 1.14 |
| Day 1 | 244 (100.0) | 13.6 (0.4) | **<0.001** | 2.14 | **7.5 (0.2)** | 0.000 | 2.74 | **4.8 (0.2)** | 0.000 | 1.63 | **1.6 (0.1)** | 0.000 | 0.85 |
| Day 2 | 201 (82.4) | 7.5 (0.4) | **<0.001** | 1.38 | **5.2 (0.2)** | 0.000 | 1.76 | **2.0 (0.2)** | 0.000 | 0.85 | **0.6 (0.1)** | 0.000 | 0.49 |
| Day 3 | 204 (83.6) | 5.6 (0.3) | **<0.001** | 1.15 | **4.1 (0.2)** | 0.000 | 1.41 | **1.3 (0.1)** | 0.000 | 0.65 | **0.4 (0.1)** | 0.000 | 0.47 |
| Day 4 | 197 (80.7) | 4.3 (0.3) | **<0.001** | 1.07 | **3.4 (0.2)** | 0.000 | 1.29 | **0.9 (0.1)** | 0.000 | 0.51 | 0.3 (0.1) | 0.001 | 0.47 |
| Day 5 | 190 (77.9) | 3.5 (0.3) | **<0.001** | 0.93 | **2.8 (0.2)** | 0.000 | 1.10 | **0.6 (0.1)** | 0.000 | 0.42 | 0.28 (0.10) | 0.005 | 0.42 |
| Day 6 | 190 (77.9) | 3.2 (0.3) | **<0.001** | 0.84 | **2.5 (0.2)** | 0.000 | 0.95 | **0.5 (0.1)** | 0.000 | 0.39 | 0.25 (0.10) | 0.013 | 0.43 |
| Day 7 | 194 (79.5) | 2.4 (0.3) | **<0.001** | 0.68 | **2.0 (0.2)** | 0.000 | 0.78 | **0.4 (0.1)** | 0.000 | 0.30 | 0.20 (0.10) | 0.043 | 0.43 |
| Day 10 | 178 (73.0) | 2.3 (0.3) | **<0.001** | 0.68 | **1.7 (0.2)** | 0.000 | 0.72 | **0.5 (0.1)** | 0.000 | 0.38 | 0.2 (0.1) | 0.033 | 0.45 |
| Day 14 | 176 (72.1) | 1.7 (0.2) | **<0.001** | 0.56 | **1.3 (0.2)** | 0.000 | 0.58 | **0.3 (0.1)** | 0.000 | 0.29 | 0.2 (0.1) | 0.032 | 0.48 |
| Week 4 | 170 (69.7) | 1.0 (0.2) | **<0.001** | 0.34 | **0.8 (0.2)** | 0.000 | 0.34 | **0.2 (0.1)** | 0.011 | 0.20 | 0.2 (0.1) | 0.055 | 0.52 |
| **Adults <50 years** | | | | | | | | | | | | | |
| Prior to infection^c^ | 364 (100.0) | 1.5 (1.9) |  |  | 1.4 (1.7) | . |  | 0.1 (0.4) | . |  | 0.0 (0.1) | . |  |
| Worst Moment | 364 (100.0) | **18.3 (0.3)** | **<0.001** | 2.97 | **8.4 (0.1)** | 0.000 | 3.39 | **6.8 (0.1)** | 0.000 | 2.75 | **3.4 (0.1)** | 0.000 | 1.28 |
| Day 1 | 364 (100.0) | **14.7 (0.3)** | **<0.001** | 2.31 | **7.7 (0.1)** | 0.000 | 2.73 | **5.3 (0.1)** | 0.000 | 1.92 | **2.0 (0.1)** | 0.000 | 1.00 |
| Day 2 | 307 (84.3) | **7.8 (0.3)** | **<0.001** | 1.48 | **5.1 (0.2)** | 0.000 | 1.76 | **2.2 (0.1)** | 0.000 | 0.98 | **0.6 (0.1)** | 0.000 | 0.54 |
| Day 3 | 312 (85.7) | **5.8 (0.3)** | **<0.001** | 1.27 | **4.0 (0.2)** | 0.000 | 1.49 | **1.5 (0.1)** | 0.000 | 0.77 | **0.5 (0.1)** | 0.000 | 0.54 |
| Day 4 | 306 (84.1) | **4.4 (0.2)** | **<0.001** | 1.13 | **3.3 (0.1)** | 0.000 | 1.25 | **1.0 (0.1)** | 0.000 | 0.65 | **0.3 (0.1)** | 0.000 | 0.43 |
| Day 5 | 284 (78.0) | **3.2 (0.2)** | **<0.001** | 0.91 | **2.5 (0.1)** | 0.000 | 0.98 | **0.7 (0.1)** | 0.000 | 0.50 | 0.2 (0.1) | 0.012 | 0.38 |
| Day 6 | 293 (80.5) | **2.8 (0.2)** | **<0.001** | 0.80 | **2.1 (0.1)** | 0.000 | 0.83 | **0.6 (0.1)** | 0.000 | 0.48 | 0.2 (0.1) | 0.019 | 0.36 |
| Day 7 | 278 (76.4) | **2.3 (0.2)** | **<0.001** | 0.69 | **1.7 (0.1)** | 0.000 | 0.74 | **0.5 (0.1)** | 0.000 | 0.42 | 0.2 (0.1) | 0.024 | 0.35 |
| Day 10 | 261 (71.7) | **1.7 (0.2)** | **<0.001** | 0.62 | **1.3 (0.1)** | 0.000 | 0.63 | **0.4 (0.1)** | 0.000 | 0.37 | 0.1 (0.1) | 0.138 | 0.37 |
| Day 14 | 256 (70.3) | **1.2 (0.2)** | **<0.001** | 0.43 | **0.8 (0.1)** | 0.000 | 0.40 | **0.3 (0.1)** | 0.000 | 0.32 | 0.1 (0.1) | 0.091 | 0.45 |
| Week 4 | 256 (70.3) | **0.6 (0.2)** | **<0.001** | 0.27 | **0.4 (0.1)** | 0.003 | 0.20 | **0.2 (0.0)** | 0.000 | 0.28 | 0.1 (0.1) | 0.181 | 0.45 |

LSE=least-square estimate; SE=standard error; ES=effect size.

^a^ Least-square estimates based on mixed models for repeated measures, which include covariates of time, high-risk status, self-reported sex, age group (≥50 vs <50), race/ethnicity, US region, social vulnerability index category, baseline outcome score, insurance type, number of comorbidities, and interaction terms between time and high-risk status, as well as time and age group, with unstructured correlation matrix for repeated measures.

^b^ ES refers to within-cohort effect size, was calculated as the least square estimate of mean change scores divided by the observed standard deviation of change scores from prior to infection to follow-up.

^c^ Mean (standard deviation) of scores prior to infection reflect observed values and were not derived using the least squares model estimates.

Bolded values indicate P<0.05 which is considered statistically significant.

**Table S8**. Prevalence of self-reported acute symptoms by severity from pre-infection to follow-up over a 1-month period among adults infected with COVID-19 aged <50 years

| **Acute symptoms** | **Time of Survey, %** | | | | | | | | | | | |
| --- | --- | --- | --- | --- | --- | --- | --- | --- | --- | --- | --- | --- |
|  | **Prior to infection**  **(N=364)** | **Worst moment**  **(N=364)** | **Day 1**  **(N=364)** | **Day 2**  **(N=307)** | **Day 3 (N=312)** | **Day 4**  **(N=306)** | **Day 5**  **(N=284)** | **Day 6**  **(N=293)** | **Day 7**  **(N=278)** | **Day 10**  **(N=261)** | **Day 14**  **(N=256)** | **Week 4**  **(N=256)** |
| **Mild to severe, %** |  |  |  |  |  |  |  |  |  |  |  |  |
| stuffy or runny nose | 23.9 | 99.2 | 97.5 | 90.2 | 82.1 | 79.7 | 70.4 | 65.5 | 60.1 | 55.6 | 43.0 | 33.6 |
| cough | 7.4 | 93.4 | 93.1 | 87.6 | 78.9 | 73.9 | 63.4 | 57.3 | 54.3 | 50.2 | 41.4 | 26.6 |
| fatigue or tiredness | 27.8 | 94.8 | 93.4 | 82.1 | 75.3 | 63.1 | 54.6 | 51.5 | 45.7 | 40.2 | 31.6 | 25.8 |
| headache | 27.5 | 91.8 | 87.4 | 61.9 | 51.3 | 43.8 | 35.6 | 33.1 | 27.3 | 22.2 | 19.5 | 21.9 |
| body aches | 19.2 | 90.9 | 81.9 | 56.0 | 41.7 | 33.7 | 29.9 | 28.0 | 25.9 | 20.7 | 18.4 | 16.0 |
| feeling hot or feverish | 3.3 | 89.3 | 76.9 | 34.2 | 22.1 | 14.4 | 8.5 | 7.2 | 5.4 | 3.1 | 2.7 | 3.5 |
| chills or shivering | 2.5 | 81.9 | 65.4 | 22.8 | 14.1 | 8.8 | 5.3 | 4.1 | 4.3 | 4.2 | 2.0 | 2.0 |
| sore throat | 8.2 | 91.5 | 83.8 | 55.7 | 46.2 | 35.3 | 26.4 | 24.2 | 19.1 | 13.8 | 10.2 | 7.0 |
| difficulty breathing | 3.3 | 58.0 | 56.0 | 38.8 | 31.7 | 24.5 | 19.0 | 18.4 | 16.9 | 14.6 | 11.3 | 7.8 |
| nausea | 2.5 | 42.9 | 37.1 | 18.9 | 13.8 | 13.1 | 8.1 | 9.6 | 6.5 | 3.8 | 3.5 | 2.3 |
| vomit | 1.4 | 11.5 | 8.0 | 2.6 | 1.6 | 0.7 | 1.4 | 1.0 | 1.4 | 0.4 | 0.8 | 1.6 |
| diarrhea | 7.4 | 36.8 | 34.3 | 26.1 | 26.9 | 21.6 | 16.6 | 14.7 | 14.4 | 12.3 | 11.3 | 9.8 |
| change of smell | 0.8 | 36.5 | 33.8 | 25.4 | 23.7 | 19.9 | 15.9 | 12.6 | 11.2 | 6.5 | 3.5 | 1.6 |
| change of taste | 1.1 | 46.7 | 42.9 | 34.2 | 26.9 | 24.2 | 18.3 | 13.0 | 9.0 | 6.1 | 3.5 | 1.6 |
| **Moderate to severe, %** |  |  |  |  |  |  |  |  |  |  |  |  |
| stuffy or runny nose | 1.7 | 85.2 | 74.2 | 40.4 | 25.0 | 19.6 | 10.9 | 11.3 | 9.4 | 6.9 | 5.9 | 2.7 |
| cough | 0.0 | 70.1 | 59.6 | 37.1 | 25.6 | 18.6 | 11.6 | 12.3 | 9.7 | 9.6 | 7.0 | 3.9 |
| fatigue or tiredness | 1.9 | 84.1 | 75.8 | 42.7 | 29.2 | 21.6 | 17.3 | 13.0 | 11.9 | 7.7 | 5.5 | 6.3 |
| headache | 1.1 | 74.5 | 62.1 | 25.4 | 18.0 | 10.8 | 9.5 | 6.1 | 8.3 | 5.4 | 5.5 | 3.5 |
| body aches | 1.1 | 73.6 | 53.6 | 16.3 | 11.5 | 7.2 | 7.8 | 7.2 | 6.8 | 3.1 | 4.7 | 3.5 |
| feeling hot or feverish | 0.3 | 70.3 | 48.9 | 10.4 | 6.7 | 2.3 | 1.8 | 2.4 | 1.1 | 1.2 | 0.8 | 0.4 |
| chills or shivering | 0.0 | 67.9 | 39.0 | 6.2 | 2.6 | 1.6 | 0.7 | 1.4 | 1.4 | 0.8 | 0.0 | 0.4 |
| sore throat | 0.6 | 74.7 | 54.1 | 18.9 | 9.3 | 2.9 | 1.8 | 1.4 | 1.1 | 2.3 | 2.0 | 2.0 |
| difficulty breathing | 0.3 | 26.4 | 21.7 | 10.4 | 8.7 | 3.9 | 1.8 | 1.7 | 1.1 | 0.8 | 1.6 | 1.2 |
| nausea | 0.3 | 17.9 | 10.4 | 2.0 | 2.6 | 2.3 | 2.1 | 2.1 | 1.8 | 0.0 | 0.4 | 0.4 |
| vomit | 0.0 | 2.8 | 0.8 | 0.3 | 0.0 | 0.0 | 0.0 | 0.0 | 0.7 | 0.0 | 0.0 | 0.0 |
| diarrhea | 0.8 | 14.0 | 12.9 | 4.9 | 5.8 | 3.3 | 3.5 | 3.1 | 2.5 | 3.1 | 1.2 | 0.8 |
| change of smell | 0.8 | 14.0 | 12.1 | 11.1 | 10.3 | 7.5 | 4.9 | 4.1 | 3.6 | 1.2 | 0.4 | 0.4 |
| change of taste | 0.3 | 9.6 | 7.1 | 8.1 | 6.7 | 5.6 | 4.6 | 2.4 | 2.2 | 0.8 | 0.4 | 0.4 |
| **Severe, %** |  |  |  |  |  |  |  |  |  |  |  |  |
| stuffy or runny nose | 0.3 | 45.3 | 26.7 | 9.5 | 5.8 | 2.3 | 1.1 | 0.7 | 0.7 | 0.4 | 0.4 | 0.4 |
| cough | 0.0 | 27.5 | 17.9 | 7.2 | 4.8 | 2.3 | 1.4 | 1.4 | 1.1 | 0.4 | 0.4 | 0.4 |
| fatigue or tiredness | 0.0 | 48.6 | 35.7 | 11.4 | 5.5 | 2.6 | 1.4 | 1.0 | 0.7 | 0.4 | 1.6 | 0.8 |
| headache | 0.0 | 39.0 | 22.8 | 6.5 | 1.9 | 2.0 | 0.4 | 1.0 | 1.4 | 1.2 | 2.0 | 1.2 |
| body aches | 0.0 | 37.1 | 20.1 | 2.9 | 1.9 | 1.0 | 0.7 | 0.7 | 0.0 | 0.8 | 1.2 | 0.0 |
| feeling hot or feverish | 0.0 | 33.2 | 15.1 | 1.0 | 1.0 | 0.0 | 0.4 | 0.7 | 0.4 | 0.4 | 0.0 | 0.0 |
| chills or shivering | 0.0 | 31.9 | 13.5 | 1.6 | 0.3 | 0.0 | 0.0 | 0.3 | 0.4 | 0.4 | 0.0 | 0.0 |
| sore throat | 0.0 | 35.4 | 20.1 | 2.9 | 1.3 | 0.3 | 0.0 | 0.3 | 0.0 | 0.0 | 0.4 | 0.8 |
| difficulty breathing | 0.0 | 7.1 | 3.0 | 0.3 | 0.3 | 0.3 | 0.0 | 0.0 | 0.0 | 0.0 | 0.0 | 0.0 |
| nausea | 0.0 | 4.7 | 1.7 | 0.0 | 1.0 | 0.0 | 0.4 | 0.3 | 0.7 | 0.0 | 0.0 | 0.0 |
| vomit | 0.0 | 1.1 | 0.3 | 0.0 | 0.0 | 0.0 | 0.0 | 0.0 | 0.0 | 0.0 | 0.0 | 0.0 |
| diarrhea | 0.3 | 4.7 | 3.0 | 1.3 | 2.2 | 0.7 | 0.4 | 1.4 | 0.7 | 0.0 | 0.4 | 0.0 |
| change of smell | 0.8 | 14.0 | 12.1 | 11.0 | 10.3 | 7.5 | 4.9 | 4.1 | 3.6 | 1.2 | 0.4 | 0.4 |
| change of taste | 0.3 | 9.6 | 7.1 | 8.1 | 6.7 | 5.5 | 4.6 | 2.4 | 2.2 | 0.8 | 0.4 | 0.4 |

**Table S9**. Summary of observed mean symptom outcome scores, score change, and effect size from pre-infection to follow-up over a 1-month period among adults infected with COVID-19 in total population and high-risk group*.

|  | **Total population** | | | | **COVID-19 high-risk group** | | | |
| --- | --- | --- | --- | --- | --- | --- | --- | --- |
|  | **n** | **Score**  **Mean (SD)** | **Score change**  **Mean (SD)** | **Effect Size** | **n** | **Score**  **Mean (SD)** | **Score change**  **Mean (SD)** | **Effect Size** |
| **Composite acute symptom score** | | | | | | | | |
| Prior to infection | 608 | 1.5 (1.9) | . (.) | . | 379 | 1.8 (2.0) | . (.) | . |
| Worst Moment | 608 | 19.0 (6.3) | 17.5 (6.2) | 2.82 | 379 | 19.3 (6.5) | 17.5 (6.3) | 2.77 |
| Day 1 | 608 | 15.6 (6.5) | 14.1 (6.4) | 2.21 | 379 | 15.8 (6.6) | 14.0 (6.5) | 2.16 |
| Day 2 | 508 | 9.1 (5.6) | 7.7 (5.3) | 1.45 | 321 | 9.5 (5.9) | 7.8 (5.6) | 1.41 |
| Day 3 | 516 | 7.2 (5.0) | 5.7 (4.7) | 1.22 | 327 | 7.6 (5.1) | 5.9 (4.8) | 1.23 |
| Day 4 | 503 | 5.8 (4.2) | 4.4 (3.9) | 1.11 | 319 | 6.3 (4.5) | 4.5 (4.1) | 1.10 |
| Day 5 | 474 | 4.7 (3.7) | 3.2 (3.6) | 0.90 | 308 | 5.1 (4.0) | 3.4 (3.8) | 0.89 |
| Day 6 | 483 | 4.4 (3.7) | 2.9 (3.6) | 0.81 | 310 | 4.7 (4.0) | 3.0 (3.8) | 0.80 |
| Day 7 | 472 | 3.8 (3.6) | 2.3 (3.4) | 0.68 | 303 | 4.3 (3.9) | 2.6 (3.7) | 0.70 |
| Day 10 | 439 | 3.3 (3.4) | 1.9 (3.1) | 0.63 | 285 | 3.7 (3.6) | 2.1 (3.3) | 0.63 |
| Day 14 | 432 | 2.7 (3.2) | 1.3 (2.9) | 0.46 | 283 | 3.0 (3.3) | 1.4 (3.0) | 0.46 |
| Week 4 | 426 | 2.1 (2.7) | 0.7 (2.6) | 0.28 | 278 | 2.5 (2.9) | 0.8 (2.8) | 0.30 |
| **Number of mild-to-severe acute symptoms** | | | | | | | | |
| Prior to infection | 608 | 1.4 (1.6) |  |  |  | 1.6 (1.7) |  |  |
| Worst Moment | 608 | 9.5 (2.3) | 8.1 (2.4) | 3.33 | 379 | 9.6 (2.3) | 8.0 (2.5) | 3.17 |
| Day 1 | 608 | 8.8 (2.6) | 7.4 (2.8) | 2.68 | 379 | 8.9 (2.7) | 7.3 (2.9) | 2.54 |
| Day 2 | 508 | 6.4 (2.9) | 5.1 (2.9) | 1.74 | 321 | 6.6 (3.0) | 5.0 (2.9) | 1.71 |
| Day 3 | 516 | 5.4 (2.9) | 4.0 (2.8) | 1.46 | 327 | 5.7 (3.0) | 4.1 (2.8) | 1.46 |
| Day 4 | 503 | 4.6 (2.7) | 3.3 (2.6) | 1.25 | 319 | 4.9 (2.8) | 3.3 (2.7) | 1.24 |
| Day 5 | 474 | 3.9 (2.5) | 2.5 (2.5) | 0.99 | 308 | 4.1 (2.6) | 2.5 (2.5) | 0.99 |
| Day 6 | 483 | 3.6 (2.6) | 2.3 (2.6) | 0.87 | 310 | 3.8 (2.7) | 2.3 (2.6) | 0.86 |
| Day 7 | 472 | 3.2 (2.5) | 1.8 (2.4) | 0.75 | 303 | 3.5 (2.6) | 1.9 (2.5) | 0.76 |
| Day 10 | 439 | 2.8 (2.4) | 1.5 (2.3) | 0.65 | 285 | 3.0 (2.5) | 1.5 (2.4) | 0.64 |
| Day 14 | 432 | 2.3 (2.2) | 1.0 (2.1) | 0.45 | 283 | 2.5 (2.3) | 1.0 (2.2) | 0.44 |
| Week 4 | 426 | 1.8 (2.1) | 0.5 (2.1) | 0.25 | 278 | 2.1 (2.2) | 0.6 (2.1) | 0.27 |
| **Number of moderate-to-severe acute symptoms** | | | | | | | | |
| Prior to infection | 608 | 0.1 (0.4) |  |  |  | 0.1 (0.5) |  |  |
| Worst Moment | 608 | 6.6 (2.6) | 6.5 (2.6) | 2.50 | 379 | 6.7 (2.6) | 6.6 (2.6) | 2.50 |
| Day 1 | 608 | 5.2 (2.8) | 5.0 (2.8) | 1.79 | 379 | 5.2 (2.9) | 5.1 (2.9) | 1.77 |
| Day 2 | 508 | 2.3 (2.4) | 2.2 (2.3) | 0.95 | 321 | 2.5 (2.5) | 2.3 (2.4) | 0.96 |
| Day 3 | 516 | 1.6 (2.0) | 1.5 (2.0) | 0.73 | 327 | 1.7 (2.1) | 1.6 (2.0) | 0.77 |
| Day 4 | 503 | 1.1 (1.7) | 1.0 (1.6) | 0.61 | 319 | 1.2 (1.8) | 1.1 (1.7) | 0.65 |
| Day 5 | 474 | 0.8 (1.4) | 0.7 (1.4) | 0.47 | 308 | 0.9 (1.5) | 0.8 (1.5) | 0.50 |
| Day 6 | 483 | 0.7 (1.3) | 0.6 (1.3) | 0.45 | 310 | 0.8 (1.5) | 0.7 (1.4) | 0.47 |
| Day 7 | 472 | 0.6 (1.3) | 0.5 (1.2) | 0.39 | 303 | 0.7 (1.4) | 0.6 (1.4) | 0.45 |
| Day 10 | 439 | 0.5 (1.2) | 0.4 (1.1) | 0.38 | 285 | 0.6 (1.2) | 0.5 (1.2) | 0.43 |
| Day 14 | 432 | 0.4 (1.0) | 0.3 (1.0) | 0.30 | 283 | 0.5 (1.1) | 0.3 (1.0) | 0.34 |
| Week 4 | 426 | 0.3 (0.8) | 0.2 (0.7) | 0.24 | 278 | 0.4 (0.8) | 0.2 (0.8) | 0.27 |
| **Number of severe acute symptoms** | | | | | | | | |
| Prior to infection |  | 0.0 (0.1) |  |  |  | 0.0 (0.1) |  |  |
| Worst Moment | 608 | 3.1 (2.6) | 3.1 (2.6) | 1.20 | 379 | 3.2 (2.6) | 3.2 (2.6) | 1.23 |
| Day 1 | 608 | 1.8 (2.0) | 1.8 (2.0) | 0.91 | 379 | 1.9 (2.1) | 1.9 (2.1) | 0.91 |
| Day 2 | 511 | 0.6 (1.2) | 0.6 (1.2) | 0.50 | 323 | 0.7 (1.3) | 0.7 (1.3) | 0.54 |
| Day 3 | 516 | 0.4 (0.9) | 0.4 (0.8) | 0.45 | 327 | 0.4 (0.9) | 0.4 (0.9) | 0.46 |
| Day 4 | 505 | 0.3 (0.7) | 0.2 (0.7) | 0.36 | 321 | 0.3 (0.7) | 0.3 (0.7) | 0.38 |
| Day 5 | 475 | 0.2 (0.6) | 0.2 (0.6) | 0.28 | 309 | 0.2 (0.7) | 0.2 (0.7) | 0.31 |
| Day 6 | 484 | 0.2 (0.5) | 0.1 (0.5) | 0.27 | 311 | 0.2 (0.6) | 0.2 (0.6) | 0.28 |
| Day 7 | 473 | 0.1 (0.5) | 0.1 (0.5) | 0.22 | 304 | 0.2 (0.6) | 0.1 (0.6) | 0.25 |
| Day 10 | 440 | 0.1 (0.4) | 0.1 (0.4) | 0.20 | 285 | 0.1 (0.4) | 0.1 (0.5) | 0.23 |
| Day 14 | 432 | 0.1 (0.4) | 0.1 (0.4) | 0.22 | 283 | 0.1 (0.4) | 0.1 (0.4) | 0.23 |
| Week 4 | 426 | 0.1 (0.3) | 0.1 (0.3) | 0.18 | 278 | 0.1 (0.3) | 0.1 (0.4) | 0.18 |

*Defined as adults aged ≥65 years or adults <65 years with ≥1 CDC-recognized underlying health condition associated with increased risk of COVID-19-related complications

**Table S10**. Summary of observed mean symptom outcome scores, score change, and effect size from pre-infection to follow-up over a 1-month period among adults infected with COVID-19 in adults ≥50 years and <50 years.

|  | **Adults≥50 years** | | | | **Adults<50 years** | | | |
| --- | --- | --- | --- | --- | --- | --- | --- | --- |
|  | **n** | **Score**  **Mean (SD)** | **Score change**  **Mean (SD)** | **ES** | **n** | **Score**  **Mean (SD)** | **Score change**  **Mean (SD)** | **ES** |
| **Composite acute symptom score** | | | | | | | | |
| Prior to infection | 244 | 1.5 (1.9) | . (.) | . | 364 | 1.5 (1.9) | . (.) | . |
| Worst Moment | 244 | 18.1 (6.5) | 16.5 (6.2) | 2.68 | 364 | 19.7 (6.2) | 18.2 (6.2) | 2.95 |
| Day 1 | 244 | 15.0 (6.5) | 13.5 (6.4) | 2.12 | 364 | 16.0 (6.4) | 14.6 (6.4) | 2.29 |
| Day 2 | 201 | 9.1 (5.7) | 7.7 (5.4) | 1.42 | 307 | 9.2 (5.5) | 7.7 (5.3) | 1.47 |
| Day 3 | 204 | 7.1 (5.1) | 5.7 (4.9) | 1.17 | 312 | 7.2 (4.9) | 5.8 (4.5) | 1.27 |
| Day 4 | 197 | 6.0 (4.3) | 4.5 (4.1) | 1.10 | 306 | 5.8 (4.1) | 4.3 (3.9) | 1.10 |
| Day 5 | 190 | 5.0 (3.8) | 3.5 (3.7) | 0.94 | 284 | 4.6 (3.7) | 3.1 (3.5) | 0.87 |
| Day 6 | 190 | 4.7 (3.8) | 3.3 (3.8) | 0.87 | 293 | 4.2 (3.7) | 2.7 (3.5) | 0.78 |
| Day 7 | 194 | 3.9 (3.5) | 2.5 (3.5) | 0.72 | 278 | 3.7 (3.6) | 2.2 (3.4) | 0.66 |
| Day 10 | 178 | 3.8 (3.6) | 2.4 (3.4) | 0.72 | 261 | 3.0 (3.2) | 1.6 (2.8) | 0.57 |
| Day 14 | 176 | 3.2 (3.2) | 1.7 (3.0) | 0.59 | 256 | 2.4 (3.1) | 1.0 (2.8) | 0.37 |
| Week 4 | 170 | 2.5 (2.9) | 1.1 (3.0) | 0.38 | 256 | 1.9 (2.6) | 0.5 (2.3) | 0.21 |
| **Number of mild-to-severe acute symptoms** | | | | | | | | |
| Prior to infection | 244 | 1.4 (1.6) |  |  | 364 | 1.4 (1.7) |  |  |
| Worst Moment | 244 | 9.2 (2.3) | 7.9 (2.4) | 3.33 | 364 | 9.7 (2.2) | 8.3 (2.5) | 3.35 |
| Day 1 | 244 | 8.7 (2.6) | 7.3 (2.7) | 2.67 | 364 | 8.9 (2.5) | 7.6 (2.8) | 2.69 |
| Day 2 | 201 | 6.5 (3.0) | 5.2 (2.9) | 1.77 | 307 | 6.4 (2.9) | 5.0 (2.9) | 1.72 |
| Day 3 | 204 | 5.5 (3.0) | 4.2 (2.9) | 1.43 | 312 | 5.4 (2.8) | 4.0 (2.7) | 1.48 |
| Day 4 | 197 | 4.8 (2.7) | 3.4 (2.6) | 1.31 | 306 | 4.6 (2.6) | 3.2 (2.6) | 1.22 |
| Day 5 | 190 | 4.1 (2.6) | 2.8 (2.5) | 1.10 | 284 | 3.7 (2.5) | 2.3 (2.5) | 0.93 |
| Day 6 | 190 | 3.9 (2.6) | 2.6 (2.7) | 0.99 | 293 | 3.4 (2.5) | 2.0 (2.5) | 0.80 |
| Day 7 | 194 | 3.4 (2.5) | 2.0 (2.5) | 0.82 | 278 | 3.0 (2.4) | 1.6 (2.3) | 0.70 |
| Day 10 | 178 | 3.1 (2.5) | 1.9 (2.4) | 0.77 | 261 | 2.5 (2.3) | 1.2 (2.1) | 0.57 |
| Day 14 | 176 | 2.7 (2.3) | 1.4 (2.2) | 0.61 | 256 | 2.0 (2.2) | 0.7 (2.0) | 0.34 |
| Week 4 | 170 | 2.2 (2.3) | 0.9 (2.3) | 0.39 | 256 | 1.6 (1.9) | 0.3 (1.9) | 0.15 |
| Prior to infection | 364 | 0.1 (0.5) |  |  | 364 | 0.1 (0.4) |  |  |
| Worst Moment | 244 | 6.3 (2.8) | 6.2 (2.8) | 2.22 | 364 | 6.8 (2.4) | 6.8 (2.5) | 2.75 |
| Day 1 | 244 | 4.9 (3.0) | 4.8 (2.9) | 1.63 | 364 | 5.3 (2.7) | 5.2 (2.7) | 1.91 |
| Day 2 | 201 | 2.3 (2.4) | 2.1 (2.4) | 0.89 | 307 | 2.3 (2.4) | 2.3 (2.3) | 0.99 |
| Day 3 | 204 | 1.5 (2.0) | 1.3 (2.0) | 0.67 | 312 | 1.6 (2.0) | 1.5 (2.0) | 0.78 |
| Day 4 | 197 | 1.1 (1.8) | 0.9 (1.7) | 0.55 | 306 | 1.1 (1.6) | 1.0 (1.5) | 0.65 |
| Day 5 | 190 | 0.7 (1.4) | 0.6 (1.4) | 0.43 | 284 | 0.8 (1.4) | 0.7 (1.4) | 0.50 |
| Day 6 | 190 | 0.7 (1.4) | 0.6 (1.4) | 0.41 | 293 | 0.7 (1.3) | 0.6 (1.2) | 0.48 |
| Day 7 | 194 | 0.6 (1.2) | 0.4 (1.3) | 0.34 | 278 | 0.6 (1.3) | 0.5 (1.2) | 0.42 |
| Day 10 | 178 | 0.7 (1.3) | 0.5 (1.3) | 0.41 | 261 | 0.4 (1.1) | 0.3 (1.0) | 0.35 |
| Day 14 | 176 | 0.5 (1.1) | 0.3 (1.1) | 0.31 | 256 | 0.4 (1.0) | 0.3 (0.9) | 0.30 |
| Week 4 | 170 | 0.3 (0.8) | 0.2 (0.8) | 0.22 | 256 | 0.3 (0.8) | 0.2 (0.7) | 0.25 |
| **Number of severe acute symptoms** | | | | | | | | |
| Prior to infection | 244 | 0.0 (0.1) |  |  | 364 | 0.0 (0.1) |  |  |
| Worst Moment | 244 | 2.7 (2.4) | 2.7 (2.4) | 1.13 | 364 | 3.4 (2.7) | 3.4 (2.7) | 1.26 |
| Day 1 | 244 | 1.6 (1.9) | 1.6 (1.9) | 0.83 | 364 | 2.0 (2.0) | 2.0 (2.0) | 0.97 |
| Day 2 | 202 | 0.6 (1.2) | 0.5 (1.2) | 0.48 | 309 | 0.6 (1.2) | 0.6 (1.2) | 0.52 |
| Day 3 | 204 | 0.3 (0.8) | 0.3 (0.8) | 0.39 | 312 | 0.4 (0.9) | 0.4 (0.8) | 0.49 |
| Day 4 | 198 | 0.3 (0.7) | 0.3 (0.7) | 0.37 | 307 | 0.2 (0.7) | 0.2 (0.6) | 0.36 |
| Day 5 | 190 | 0.2 (0.7) | 0.2 (0.7) | 0.30 | 285 | 0.2 (0.5) | 0.1 (0.5) | 0.27 |
| Day 6 | 191 | 0.2 (0.6) | 0.2 (0.6) | 0.30 | 293 | 0.1 (0.5) | 0.1 (0.5) | 0.25 |
| Day 7 | 194 | 0.1 (0.5) | 0.1 (0.5) | 0.25 | 279 | 0.1 (0.5) | 0.1 (0.5) | 0.20 |
| Day 10 | 178 | 0.1 (0.5) | 0.1 (0.5) | 0.28 | 262 | 0.1 (0.3) | 0.0 (0.3) | 0.12 |
| Day 14 | 176 | 0.1 (0.5) | 0.1 (0.5) | 0.27 | 256 | 0.1 (0.3) | 0.1 (0.3) | 0.17 |
| Week 4 | 170 | 0.1 (0.4) | 0.1 (0.4) | 0.26 | 256 | 0.0 (0.3) | 0.0 (0.2) | 0.10 |

**Table S11.** Comparisons of adults included and not included in the final analysis

| **Characteristic** | **Included (N=608)** | **Not Included (N=29483)** | **P value** |
| --- | --- | --- | --- |
| Age, years, mean (SD) | 45.9 (14.2) | 45.2 (16.8) | 0.316 |
| Sex, n (%) |  |  | **<0.0001** |
| Female | 465 (76.5) | 18,579 (63.0) |  |
| Race/Ethnicity, n (%) |  |  | **<0.0001** |
| White or Caucasian | 407 (66.9) | 16,391 (55.6) |  |
| Black or African American | 49 (8.1) | 3,118 (10.6) |  |
| Hispanic | 75 (12.3) | 4,538 (15.4) |  |
| Asian | 26 (4.3) | 1,755 (6.0) |  |
| Other ^a^ | 51 (8.4) | 3,681 (12.5) |  |
| US Region, n (%) |  |  | 0.894 |
| Northeast | 73 (12.0) | 3,188 (10.8) |  |
| South | 368 (60.5) | 18,179 (61.7) |  |
| Midwest | 133 (21.9) | 6,375 (21.6) |  |
| West | 33 (5.4) | 1,673 (5.7) |  |
| Other\Unknown | 1 (0.2) | 68 (0.2) |  |
| Insurance type, n (%) |  |  | **0.011** |
| Commercial | 480 (78.9) | 21,581 (73.2) |  |
| Government | 72 (11.8) | 4,696 (15.9) |  |
| Self-pay | 53 (8.7) | 2,749 (9.3) |  |
| Missing | 3 | 457 |  |
| Social Vulnerability Index, mean (SD) | 0.37 (0.21) | 0.27 (10.10) | 0.810 |
| **SVI category, n (%)** |  |  | 0.931 |
| <0.25 | 205 (33.7) | 9,880 (33.5) |  |
| >=0.25 and <0.5 | 244 (40.1) | 11,825 (40.1) |  |
| >=0.5 and <0.75 | 129 (21.2) | 6,074 (20.6) |  |
| >=0.75 | 30 (4.9) | 1,618 (5.5) |  |
| Missing | 0 | 86 |  |

SD=standard deviation; US=United States.

^a^ American Indian or Alaska Native, Native Hawaiian or Other Pacific Islander, Patient Refused, Other, Unknown.

^b^ Government, other, or unknown.

Note: Patient characteristics compared between those included vs. not included were retrieved from CVS Health MinuteClinic® testing database, rather than from survey. Race and ethnicity may be reported differently. Insurance type was not asked in the survey.

Bolded p-values indicate statistical significance at p <0.05

Not included patients were adults with a positive COVID-19 test contacted by email but not included in the final analytic sample, primarily due to non-engagement (n=28,533), did not meet the exclusion criteria (n=423), did not register (n=300), did not consent (n=134), and failed validity checks/did not complete first survey (n=93) (See Figure 1)

**Table S12**. Reasons for attrition

| **Reason** | **N (%)** |
| --- | --- |
| I was too busy to complete it or forgot | 68 (56.7) |
| I was sick and didn’t feel well enough to answer | 17 (14.2) |
| I had technical difficulties accessing or opening the link to the survey | 13 (10.8) |
| Other | 11 (9.2) |
| My illness-related symptoms resolved and I preferred to stop answering illness-related questions | 3 (2.5) |
| The compensation amount was not enough | 3 (2.5) |
| I get too many emails or surveys | 2 (1.7) |
| The questions were too personal or too sensitive | 1 (0.8) |
| I was too busy to complete it or forgot | 1 (0.8) |
| I changed my mind about my interest in the topics of the survey | 1 (0.8) |
| **Total, N** | 120 |

STROBE Statement—Checklist of items that should be included in reports of ***cohort studies***

|  | **Item No** | **Recommendation** | **Page No** |
| --- | --- | --- | --- |
| **Title and abstract** | 1 | (*a*) Indicate the study’s design with a commonly used term in the title or the abstract | 1 |
|  |  | (*b*) Provide in the abstract an informative and balanced summary of what was done and what was found | 2-3 |
| **Introduction** | | | |
| Background/rationale | 2 | Explain the scientific background and rationale for the investigation being reported | 3-4 |
| Objectives | 3 | State specific objectives, including any prespecified hypotheses | 4 |
| **Methods** | | | |
| Study design | 4 | Present key elements of study design early in the paper | 4-5 |
| Setting | 5 | Describe the setting, locations, and relevant dates, including periods of recruitment, exposure, follow-up, and data collection | 4-5 |
| Participants | 6 | (*a*) Give the eligibility criteria, and the sources and methods of selection of participants. Describe methods of follow-up | 6-7 |
|  |  | (*b*) For matched studies, give matching criteria and number of exposed and unexposed | n/a |
| Variables | 7 | Clearly define all outcomes, exposures, predictors, potential confounders, and effect modifiers. Give diagnostic criteria, if applicable | 5-6 |
| Data sources/ measurement | 8* | For each variable of interest, give sources of data and details of methods of assessment (measurement). Describe comparability of assessment methods if there is more than one group | 5 |
| Bias | 9 | Describe any efforts to address potential sources of bias | 6 |
| Study size | 10 | Explain how the study size was arrived at | Figure 1 |
| Quantitative variables | 11 | Explain how quantitative variables were handled in the analyses. If applicable, describe which groupings were chosen and why | 6-7 |
| Statistical methods | 12 | (*a*) Describe all statistical methods, including those used to control for confounding | 7 |
|  |  | (*b*) Describe any methods used to examine subgroups and interactions | 7 |
|  |  | (*c*) Explain how missing data were addressed | 7 |
|  |  | (*d*) If applicable, explain how loss to follow-up was addressed | 7 |
|  |  | (*e*) Describe any sensitivity analyses | 7 |
| **Results** | | |  |
| Participants | 13* | (a) Report numbers of individuals at each stage of study—eg numbers potentially eligible, examined for eligibility, confirmed eligible, included in the study, completing follow-up, and analysed | Figure 1 |
|  |  | (b) Give reasons for non-participation at each stage | Figure 1 |
|  |  | (c) Consider use of a flow diagram | Figure 1 |
| Descriptive data | 14* | (a) Give characteristics of study participants (eg, demographic, clinical, social) and information on exposures and potential confounders | 7-8 |
|  |  | (b) Indicate number of participants with missing data for each variable of interest | Table 1 |
|  |  | (c) Summarise follow-up time (eg, average and total amount) | Tables 2 and 3 |
| Outcome data | 15* | Report numbers of outcome events or summary measures over time | 8-9 |

| Main results | 16 | (*a*) Give unadjusted estimates and, if applicable, confounder-adjusted estimates and their precision (e.g., 95% confidence interval). Make clear which confounders were adjusted for and why they were included | 8-9 |
| --- | --- | --- | --- |
|  |  | (*b*) Report category boundaries when continuous variables were categorized | Table 1 |
|  |  | (*c*) If relevant, consider translating estimates of relative risk into absolute risk for a meaningful time period |  |
| Other analyses | 17 | Report other analyses done—eg analyses of subgroups and interactions, and sensitivity analyses | 9 |
| **Discussion** | | | |
| Key results | 18 | Summarise key results with reference to study objectives | 9-10 |
| Limitations | 19 | Discuss limitations of the study, taking into account sources of potential bias or imprecision. Discuss both direction and magnitude of any potential bias | 12-13 |
| Interpretation | 20 | Give a cautious overall interpretation of results considering objectives, limitations, multiplicity of analyses, results from similar studies, and other relevant evidence | 9-14 |
| Generalisability | 21 | Discuss the generalisability (external validity) of the study results | 10-12 |
| **Other information** | | | |
| Funding | 22 | Give the source of funding and the role of the funders for the present study and, if applicable, for the original study on which the present article is based | Funding section |

*Give information separately for exposed and unexposed groups.

**Note:** An Explanation and Elaboration article discusses each checklist item and gives methodological background and published examples of transparent reporting. The STROBE checklist is best used in conjunction with this article (freely available on the Web sites of PLoS Medicine at http://www.plosmedicine.org/, Annals of Internal Medicine at http://www.annals.org/, and Epidemiology at http://www.epidem.com/). Information on the STROBE Initiative is available at http://www.strobe-statement.org.
